# MOSAIC: Explainable AI for Reproducible Histologic Grading and Prognostic Stratification in Breast Cancer

**DOI:** 10.64898/2026.03.11.26348043

**Authors:** Pranali Sonpatki, Shashank Gupta, Avishek Biswas, Sharada Patil, Shweta Tyagi, Lavanya Balakrishnan, Harnisha Mistry, Preeti Doshi, Kunda Jagadale, Parineeta Shelke, Loma Parikh, Mithun Shah, Reena Bharadwaj, Shefali Desai, Madhura Kulkarni, C B Koppiker, Jyothi Prabhu, Udayan Kachchhi, Nameeta Shah

## Abstract

Nottingham histologic grading is essential for breast cancer prognostication but suffers from inter-observer variability in assessing mitotic activity, nuclear pleomorphism, and tubule formation. We developed MOSAIC (Mammary Oncology Spatial Analysis and Intelligent Classification), an explainable AI framework designed to perform component-wise grading by independently modeling these three histologic features. Model outputs were calibrated using a two-phase pathology study to establish clinically reproducible scoring thresholds and were subsequently evaluated across public datasets and multi-institutional Indian cohorts. MOSAIC demonstrated robust performance, with AI-derived grades providing independent prognostic information (HR >= 1.8 in two datasets, p = < 0.001) and improved survival stratification compared to traditional methods. In pathologist calibration studies, AI-assisted scoring significantly reduced variability, specifically achieving near-perfect agreement in mitotic scoring with a weighted κ up to 0.98. Accuracy and Cohen’s kappa (κ) analysis further characterized the model’s technical performance across components: Tubule formation showed the highest agreement (Accuracy >= 0.6607, κ = 0.549), followed by overall Grade (Accuracy = 0.5637, κ = 0.539) and Mitotic activity (Accuracy = 0.4985, κ = 0.4), while Nuclear pleomorphism proved the most challenging (Accuracy = 0.3303, κ = 0.271). Comparative survival models confirmed that AI-derived grades were more significant predictors of risk than manual pathologist-assigned grades, with the AI model yielding a superior global p-value (5.9 × 10^-7^) and lower AIC (769.61). These results indicate that MOSAIC enables reproducible, interpretable grading by decomposing assessment into pathology-aligned components. By enhancing consistency while preserving prognostic relevance, this framework supports explainable AI as a viable assistive tool for routine breast cancer pathology.

## Introduction

Histologic tumor grading remains a cornerstone of diagnostic, prognostic assessment, and treatment planning in breast cancer, providing essential information regarding tumor aggressiveness and expected clinical behavior. The Nottingham histologic grade (NHG), derived from the evaluation of mitotic activity, nuclear pleomorphism, and tubule formation, is among the most widely adopted grading frameworks for breast cancer in routine pathology practice. Despite its established clinical value, NHG assessment is inherently subjective and labor-intensive, with substantial inter- and intra-observer variability, particularly for mitotic activity and nuclear pleomorphism.^1^ Such variability can result in inconsistent grade assignment and reduced diagnostic confidence, with downstream effects such as the potential for over- or under-treatment in patients with discordant grades.

The increasing adoption of whole-slide imaging has facilitated the application of artificial intelligence (AI) to histopathologic assessment, enabling deep learning models to detect and quantify morphologic features directly from digitized slides. Prior studies have reported strong concordance between AI-derived and pathologist-assigned scores for mitotic activity and nuclear pleomorphism,^2^ as well as accurate prediction of NHG with clinically relevant prognostic associations.^3^ More recently, explainable AI (xAI) approaches have emphasized interpretability by aligning model outputs with established histologic criteria, achieving grading performance comparable to multi-institutional pathologist cohorts.^4^

Collectively, these advances highlight the potential of AI-assisted grading to improve reproducibility and efficiency in breast cancer pathology. Models such as DeepGrade and foundation-model-based approaches have demonstrated prognostic equivalence to conventional NHG while reducing inter-pathologist variability.^5,6^ However, many existing approaches rely on end-to-end grade prediction or dataset-specific optimization, limiting their utility in diverse, real-world clinical environments where stain variability, scanner heterogeneity, and institutional workflow differences are prevalent. In addition, relatively few studies incorporate systematic, pathology-driven calibration to define and refine component-level grading thresholds and evaluate performance across geographically and demographically diverse clinical cohorts.

To address these limitations, we developed MOSAIC (Mammary Oncology Spatial Analysis and Intelligent Classification), an explainable AI framework designed to perform component-wise NHG assessment from whole-slide images (WSIs). MOSAIC independently models mitotic activity, nuclear pleomorphism, and tubule formation, using scoring thresholds derived from a multi-phase pathology calibration study conducted prior to validation. In this study, we evaluate MOSAIC’s technical performance, its impact on inter-observer agreement, and its prognostic relevance across public datasets and multiple Indian clinical cohorts. By emphasizing interpretability, calibration, and multi-institutional validation, this work aims to advance AI as a pathology-aligned assistive tool for consistent, reproducible, and improved breast cancer grading.

## Materials and Methods

### Breast cancer datasets and patient cohorts

This study utilized WSIs from clinically diagnosed breast cancer cases for training and validation of the AI models. The datasets comprised both publicly available sources and data from private institutions in India.

### Ethical approval

All patient slides and associated clinical data from private institutions were collected retrospectively after obtaining approval from the respective institutional ethics committees, in accordance with the Declaration of Helsinki. Waivers of informed consent were granted where applicable due to the retrospective and de-identified nature of the data. The image and data were obtained with appropriate MTA and MoU agreements.

Ethics approvals were obtained from:

- St John’s Research Institute (SJRI), IEC/1/364/2023
- Prashanti Cancer Care Mission (PCCM), Prashanti Cancer Care Mission Independent Ethics Committee (2 November 2023); IEC project No. 0025102023
- Samved Medicare Hospital, Sangini Ethics Committee (23 September 2024)
- Dr. Udayan’s Laboratory (UL), Pranayam Lung & Heart Institute Ethics Committee (17 March 2025)
- Bharati Vidyapeeth Deemed to be University Medical College (BVDUMC), BVDUMC/IEC/215/25-26
- Zydus Hospitals, Zydus Hospital Ethics Committee (9 September, 2022)

### Training dataset

Whole Slide Image (WSIs) from histologically confirmed invasive ductal carcinoma cases were used for model training and internal development. Training data were obtained from a combination of publicly available datasets and private institutions in India.

Publicly available training datasets included subsets of The Cancer Genome Atlas breast cancer cohort (TCGA-BRCA)^7^, and the MIDOG dataset^8^. These datasets provided curated annotations of mitotic activity, nuclear pleomorphism, and tissue-level morphology. In addition, publicly available spatial transcriptomics data from the Xenium Prime FFPE Human Breast Cancer dataset (10x Genomics)^9^ were used to support cross-modal analyses linking histological features with spatially resolved gene expression patterns. Training data were also obtained from private Indian institutions, SJRI, PCCM, and UL (Table 1).

**Table 1:** Datasets and number of images used for model training and validation Clinical data and outcome availability.

| Model | Datasets | Image patches | Image patches | Image patch | Image |
| --- | --- | --- | --- | --- | --- |

**Table 1: Datasets and number of images used for model training and validation**
|  | (number of WSIs) used | used for model training | used for validation | size (pixels) | resolution |
| --- | --- | --- | --- | --- | --- |
| Segmentation | SJRI (14), TCGA-BRCA (48), TUPAC (10), UL (13) | 20,283 | 6,899 | 256 × 256 | 20x |
| Tubule formation | UL (17), TCGA-BRCA (35) | 1,688 | 593 | 256 × 256 | 20x |
| Mitotic detection | UL (1), TUPAC (1), MIDOG (378) | 17,311 | 4,659 | 384 × 384 | 40x |
| Nuclear detection | NuCLs dataset (122) SJRI (35), UL (16), PCCM (3), Xenium 10X Genomics (1) | 25,134 | 6,254 | 256 × 256 | 40x |

### Training Data Annotation

The training dataset integrated publicly available mitotic and nuclear labels from TUPAC^10^ and NuCLS^11^ (TCGA-BRCA) with high-fidelity ground truth established through independent regional and object-level annotations by two expert breast pathologists (U.K. and S.P.). Annotated WSIs were selected to represent a broad range of histologic appearances and staining patterns. Pathologist annotations were used to supervise both tissue-level segmentation and cellular-level detection tasks. For one representative case, publicly available spatial transcriptomics data provided by 10x Genomics (Xenium platform) were used as a supportive reference during annotation and were cross-verified by the pathologists. Annotation details, including the number of slides used for each model, are summarized in Table 1.

### Pathology calibration study dataset

A multi-phase pathology calibration study was conducted to derive component-level scoring thresholds for mitotic activity, nuclear pleomorphism, and tubule formation. The calibration datasets were distinct from the primary training and validation datasets.

Phase 1 of the calibration study included 30 breast cancer WSIs, comprising 17 cases from TCGA-BRCA and 13 cases from UL.

Phase 2 of the calibration study included 251 WSIs from the National Institute of Health (NIH) Prostate, Lung, Colorectal, and Ovarian Cancer Screening Trial (PLCO) dataset.^12^ Only breast cancer cases from the NIH-PLCO cohort were used for this study.

### Validation dataset

Independent validation was performed using breast cancer WSIs from publicly available datasets and multiple Indian clinical cohorts.

Public validation datasets included TCGA-BRCA (*n* = 1,020) and NIH-PLCO (*n* = 669). Indian validation cohorts included SJRI (Bangalore, Karnataka; *n* = 192), UL (Vadodara, Gujarat; *n* = 67), Samved Medicare Hospital (Ahmedabad, Gujarat; *n* = 37), BVDUMC (Pune, Maharashtra; *n* = 12), and Zydus Hospitals (Ahmedabad, Gujarat; *n* = 9).

For all private-institution validation cohorts, only WSIs from surgical specimens were included. For institutions contributing to both training and validation datasets (SJRI and UL), strict case-level separation was enforced, and only non-overlapping surgical specimens were used for validation to prevent data leakage. Sample sizes were determined by dataset availability.

Demographic and clinicopathologic variables for TCGA-BRCA and NIH-PLCO cohorts were obtained from their respective publicly available clinical data repositories and are summarized in Supplementary File 1. For private-institution cohorts, clinicopathologic data were collected retrospectively where available. Among the private-institution datasets, longitudinal outcome data were available only for SJRI cases. Accordingly, survival and prognostic analyses were restricted to the TCGA-BRCA, NIH-PLCO, and SJRI cohorts. Validation cohorts without outcome data were used exclusively for concordance and reproducibility analyses.

### Image Acquisition and Management

WSIs from SJRI were digitized using the Motic VM1 Virtual Microscopy digital slide system (Xiamen, China) and the Morphle whole slide scanner (SN_MIND202101003) (New York, USA), while slides from PCCM were digitized using an OptraSCAN 15 scanner (Pune, India). Slides from BVDUMC, Samved Medicare Hospital, UL, and Zydus Hospitals were scanned using the Motic EasyScan Pro 6 scanner (Xiamen, China).

All slides from private institutions were scanned at 40x magnification, corresponding to a spatial resolution of approximately 0.25 µm per pixel. Publicly available WSIs from TCGA and NIH-PLCO were used as provided by their respective repositories, with acquisition parameters determined by the source datasets.

All WSIs were de-identified by the host institutions and provided for analysis. The digital WSIs were securely stored on a centralized server with controlled access. No manual color correction was performed during image acquisition. Variability in staining intensity and scanner characteristics was intentionally preserved to reflect real-world clinical conditions.

### AI-Based Detection of Histologic Features

MOSAIC employs two complementary deep learning models designed for distinct but interdependent tasks: semantic segmentation and object detection. Semantic segmentation is used to identify tissue compartments and glandular structures, while object detection is used to localize and quantify individual cellular features.

### Tissue and Tubule Segmentation

Semantic segmentation was performed using a U-Net-based architecture^13^ to delineate tissue compartments and identify glandular structures relevant to tubule formation assessment. A total of 85 WSIs were used for segmentation model training. The WSIs were annotated into multiple histologic classes, including cellular tumor (CT), stroma (ST), adipose tissue (FAT), necrosis (NE), skin, normal breast tissue or ductal hyperplasia (BR-D), and regions affected by folds or other artifacts.

Tubules were defined as glandular structures with identifiable luminal formation and were annotated as a binary class (tubule vs background). The segmentation model was applied to full WSIs to generate spatial maps of detected tubules. Detected tubules were subsequently contextualized using tissue segmentation outputs. Tubules overlapping regions classified as CT were labeled as tubules within CT, while tubules located outside CT regions were labeled as tubules not within CT. Quantitative measures of tubule formation were derived from tubules within CT and normalized by tumor area for downstream scoring, as described in subsequent sections.

### Mitotic Activity Detection Model

Mitotic activity was assessed using a dedicated AI-based detection pipeline designed to identify mitotic figures within standardized high-power fields (HPFs) in WSIs. The MIDOG dataset provided annotated WSIs or regions of interest (ROIs) with ground truth labels for mitotic figures curated by experienced pathologists. The WSI from the TUPAC dataset was included as a negative sample, containing non-mitotic cells with morphological features resembling mitotic figures, to explicitly model mitotic mimetics and improve discrimination between true mitoses and non-mitotic nuclei.

The mitotic detection model was trained to classify detected objects into two categories: mitotic figures (MIT) and mitotic mimetics (MIMI). The model first identifies mitotic figures across tumor regions within the WSI. Based on the spatial distribution of detected mitotic figures, the model then selected ten consecutive HPFs with the highest mitotic activity for standardized analysis. Each HPF was equivalent to a field with a diameter of 0.51 mm (approximately 0.204 mm² area), consistent with widely accepted pathological definitions. Within the selected HPFs, object-level classification of detected nuclei into mitotic figures and mitotic mimetics was performed, generating spatially resolved detections that formed the basis for downstream mitotic activity aggregation and scoring.

### Nuclear Detection and Cell-Type Identification

A DINO-DETR-based^14^ object detection model was trained to localize nuclei and assign them to biologically relevant cell-type categories.

For model development, over 220,000 labeled nuclei from the NuCLS dataset derived from TCGA-BRCA were used. The model was trained to detect and classify nuclei into multiple cell types, including tumor cells (CE), normal epithelial cells (nCE), tumor-infiltrating lymphocytes (TIL), endothelial cells (End), fibroblasts (Fib), neutrophils (Neu), plasma cells (Plasma), and macrophages (Mph).

### Cell-Type Reference Annotation Using Spatial Transcriptomics

To support robust cell-type definition, publicly available spatial transcriptomics data from the 10x Genomics Xenium Human Breast Tissue dataset were used as a reference for cell-type annotation. Xenium transcriptomic profiles were clustered using a modified Recursive Consensus Clustering (RCC) approach.^15^ Initial (Level 1) clusters were defined, and cluster-specific marker genes were used to assign major cell types. Cell-type assignments were cross-validated using marker sets reported in prior single-cell and spatial transcriptomics studies.^16,17,18^ The Xenium dataset was used solely as a reference for cell-type definition and annotation consistency and did not serve as direct training supervision for the WSI-based detection model.

### Nuclear Pleomorphism Feature Extraction

For nuclear pleomorphism analysis, tumor cells identified as CE by the nuclei detection model were selected. For each detected tumor nucleus, the area of the predicted bounding box was computed as a quantitative proxy for nuclear size. These nucleus-level measurements were subsequently aggregated at the slide level to derive pleomorphism-related features, which were mapped to nuclear pleomorphism scores as described in later sections (Results).

### Evaluation of AI Model Performance

AI models were evaluated independently on task-specific held-out datasets using precision, recall, and F1 scores, supplemented by pixel-wise accuracy for the semantic segmentation of tissue and tubule formations. To ensure biological and clinical consistency, nuclei classification was calibrated against Xenium spatial transcriptomics, while MOSAIC outputs were evaluated for directional concordance with TCGA-BRCA molecular data, specifically ssGSEA of mitotic and cell-type specific gene sets, and established pathologist-assigned metrics from prior publications. All reported metrics reflect standalone model behavior, established prior to downstream grading or survival analyses to ensure an unbiased assessment of the framework’s diagnostic performance.

The biological accuracy of the nuclei detection and classification model was validated using three 10x Genomics Xenium spatial transcriptomics images. For each segmented cell, transcript overlaps were quantified across the 380-gene panel and aggregated by predicted cell type to assess the enrichment of lineage-specific transcripts, ensuring high-fidelity concordance between morphological classification and molecular expression.

### AI Model Performance

The technical performance of the MOSAIC framework was evaluated independently for each model component using held-out validation data (Supplementary Table S1).

For tissue segmentation, the U-Net model achieved an overall accuracy of 0.9, with weighted precision, recall, and F1-scores of 0.8988, 0.8989, and 0.8987, respectively. The tubule formation segmentation model demonstrated higher performance, with an overall accuracy of 0.97 and weighted precision, recall, and F1-scores of 0.9744, 0.9745, and 0.9744, respectively.

Mitotic activity detection using the DINO-DETR model achieved a mean average precision (mAP) of 76.1%, with a recall of 0.970 and an F1-score of 0.853, indicating high sensitivity for mitotic figure detection. Nuclear detection and classification, also performed using DINO-DETR, yielded a mAP of 70.6%, a recall of 0.895, and an F1-score of 0.789, supporting reliable localization of nuclei and extraction of quantitative nuclear features relevant to pleomorphism assessment.

Together, these results establish the technical validity of the individual MOSAIC modules and support their use in subsequent analyses evaluating pathological concordance, biological consistency, and clinical relevance.

### Pathology Calibration and Evaluation Study Design Overview

A multi-phase pathology study was conducted to evaluate AI-assisted assessment of NHG components and to establish calibrated criteria for integrating MOSAIC outputs into routine grading workflows. The study comprised two sequential phases: an initial controlled evaluation (Phase 1) followed by a large-scale assessment on an expanded dataset (Phase 2).

### Participants

Seven board-certified pathologists with 5-25 years of experience in breast pathology and Nottingham grading participated in the study. All pathologists independently assessed each case according to standard NHG criteria under predefined reading conditions.

### Phase 1: Controlled Calibration Study

#### Dataset

Phase 1 utilized a curated set of 30 digitized breast cancer WSIs, including 17 cases from the TCGA-BRCA dataset and 13 cases from UL. Cases were selected to represent a broad range of histologic patterns and grading difficulty encountered in routine practice.

#### Study Sessions

Phase 1 consisted of four grading sessions designed to isolate the effects of re-evaluation, definitional alignment, and AI assistance on component-level grading. For mitotic assessment, ten HPFs were provided for each image in all sessions.

### Session 1: Baseline Assessment (No AI)

All participating pathologists independently reviewed the full set of 30 images without AI assistance. This session served to establish baseline grading behavior and familiarize participants with the digital platform.

### Moderator-Led Calibration Session

Following Session 1, a structured calibration meeting was conducted to harmonize interpretive criteria and align definitions. Pathologists reviewed representative fields and discussed borderline cases related to mitotic figures, nuclear features, and glandular architecture, referencing established grading guidelines.

### Session 2: Repeat Assessment (No AI)

After a washout period of one month, pathologists re-evaluated the same 30 images in a randomized order, without AI assistance. This session enabled assessment of intra-observer consistency. For the mitotic component only, pathologists had access to their own and their peers’ prior annotations from Session 1.

### Session 3: AI-Assisted Assessment

In Session 3, pathologists re-evaluated the same image set, presented in a randomized order, with AI assistance enabled. AI assistance included:

- Visual overlays highlighting detected mitotic figures, nuclei, and tubules
- Identification of HPFs with high predicted mitotic activity

### Session 4: Repeat AI-Assisted Assessment

Session 4 repeated the AI-assisted evaluation with randomized image order to assess the stability of AI influence across repeated exposure. For nuclear pleomorphism, additional visualizations summarizing distributions of nuclear size, chromatin intensity, and vesicularity were provided. For mitotic assessment, pathologists had access to AI annotations and their own and peers’ annotations from Session 3.

### Derivation and Finalization of NHG Component Scoring Criteria

Following completion of the controlled calibration sessions in Phase 1 and prior to initiation of Phase 2, scoring criteria for each NHG component’s mitotic activity, nuclear pleomorphism, and tubule formation were finalized to translate continuous, AI-derived quantitative features into discrete component scores aligned with conventional NHG definitions.

Scoring criteria were defined based on a combination of (i) distributions of MOSAIC-derived quantitative features, (ii) patterns observed in pathologist-assigned component scores during Phase 1 sessions, and (iii) consensus discussions among participating pathologists regarding clinically meaningful thresholds. The objective was to establish scoring thresholds that were interpretable, reproducible, and consistent with routine pathology practice, rather than to optimize agreement with any single observer.

### Mitotic Activity Scoring

Mitotic activity scores were assigned as follows:

**Score 1:** ≤ 7 mitotic figures per 10 HPFs

**Score 2:** > 7 and ≤ 14 mitotic figures per 10 HPFs

**Score 3:** > 14 mitotic figures per 10 HPFs

In cases where ten HPFs could not be identified due to a limited tumor area, a predefined alternative criterion based on mitotic density per tumor cell count was applied. By default, such cases were assigned a mitotic score of 1. Escalation to mitotic score 3 was applied only when the mitotic density exceeded a high-confidence threshold, defined as:

**Score 3:** > 40 mitotic figures per 10,000 tumor cells

### Nuclear Pleomorphism Scoring

Nuclear pleomorphism scoring was based on the mean nuclear area and the interquartile range (IQR) of nuclear area distributions within tumor regions.

**Score 1:** Mean nuclear area < 9.5 μm²

**Score 2:** Mean nuclear area ≥ 9.5 μm² and ≤ 10 μm², or mean nuclear area ≤ 11 μm² with an IQR ≤ 4, provided the criteria for Score 1 were not met

**Score 3:** Mean nuclear area > 11 μm², or mean nuclear area > 10 μm² with an IQR > 4, provided the criteria for Score 1-2 were not met

### Tubule Formation Scoring

Tubule formation was quantified as the density of tubules within CT regions, expressed as the number of tubules per mm². Component scores were assigned as follows:

**Score 1:** > 7.5 tubules per mm²

**Score 2:** > 1.5 and ≤ 7.5 tubules per mm²

**Score 3:** ≤ 1.5 tubules per mm²

### Derivation of the Overall AI Grade

The overall AI-derived grade for each case was calculated by summing the component scores for mitotic activity, nuclear pleomorphism, and tubule formation. The total AI score was mapped to a final grade using standard Nottingham criteria:

**Grade 1:** Total AI score of 3 - 5

**Grade 2:** Total AI score of 6 - 7

**Grade 3:** Total AI score of 8 - 9

### Phase 2: Large-Cohort Evaluation Study Dataset

Phase 2 utilized 251 WSIs from the NIH-PLCO breast cancer cohort with available long-term clinical outcome data.

### Study Sessions

Phase 2 consisted of three structured reading sessions designed to assess grading behavior with and without AI assistance across a larger cohort.

### Session 1: Baseline Assessment

All pathologists independently graded a common subset of 41 images without AI assistance to establish baseline inter-observer variability.

### Session 2: Individual Repeat Assessment

Each pathologist independently graded a unique set of 30 images randomly assigned to them without AI assistance. This session served as an internal control for intra-observer consistency.

### Session 3: AI-Assisted Assessment

Pathologists reviewed 212 images with AI assistance enabled. This dataset included images previously reviewed in Sessions 1 and 2. AI support included visual overlays, component-level quantitative outputs, and suggested NHG component scores derived from model outputs, accompanied by corresponding visual and quantitative evidence. For mitotic assessment, ten HPFs were provided for each image.

For each pathologist, the time required to complete the grading of individual WSIs was recorded during both Sessions 1 and 3 using digital log timestamps within the viewing platform. Average grading time per slide and cumulative time per session were calculated and compared to quantify the effect of MOSAIC assistance on turnaround time.

### Statistical analysis

#### Concordance with Pathology Grading

Concordance between MOSAIC-derived grades and pathologist assessments was evaluated using Cohen’s kappa (κ) coefficient. For comparisons across independent validation datasets, weighted κ was used to account for the ordinal nature of NHG (Grades 1-3).

For the multi-phase pathology study (Phase 1 and Phase 2), both weighted and unweighted κ statistics were computed to comprehensively assess inter- and intra-observer agreement under different reading conditions. AI-derived grades were directly compared with pathologist-assigned grades. Confusion matrices were constructed to summarize exact grade agreement and directional grade shifts between MOSAIC predictions and pathologist assessments.

### Survival Analysis

Survival analyses were conducted using three independent cohorts: TCGA-BRCA, NIH-PLCO breast cancer cohort, and the Indian SJRI cohort. In TCGA-BRCA, overall survival (OS), disease-specific survival (DSS), disease-free interval (DFI), and progression-free interval (PFI) were analyzed using publicly available clinical annotations. In the NIH-PLCO breast cancer cohort, OS was defined as all-cause mortality and calculated as the difference between mortality_exitdays (Days from randomization until mortality exit date) and breast_exitdays (Days from trial entry randomization to cancer diagnosis for participants with breast cancer, or to trial exit otherwise), with censoring applied to individuals alive at last follow-up. In the SJRI cohort, OS was used for survival analysis. For all cohorts, survival times were converted to months.

Kaplan - Meier survival estimates were generated, and statistical differences between groups were assessed using the log-rank test, including tests for trend where appropriate. Cox proportional hazards regression models were fitted for each cohort to estimate hazard ratios, incorporating clinical covariates, including age at diagnosis, stage, and molecular subtype, where available. Proportional hazards assumptions were assessed using standard diagnostic methods.

### Optimised binary classification for survival analysis

Continuous parameters calculated by MOSAIC were converted into binary risk groups (high vs. low risk) to enable clinically interpretable survival stratification. The optimal cutoff was determined in the TCGA cohort by maximizing the log-rank test statistic across candidate thresholds. Survival differences between groups were assessed using Kaplan-Meier estimates and compared with the log-rank test. Hazard ratios (HRs) and 95% confidence intervals (CIs) were estimated using Cox proportional hazards regression.

### Single-Sample Gene Set Enrichment Analysis

Single-sample gene set enrichment analysis (ssGSEA) was performed on log2-transformed, counts-per-million (CPM) normalized TCGA transcriptomic data to explore biological correlates of AI-derived quantitative features. Gene sets corresponding to distinct cell types were curated from previously published single-cell and spatial transcriptomics studies.^16,17,18^ ssGSEA scores were used for exploratory correlation analyses and were not incorporated into model training or grading calibration.

### General Statistical Testing

Comparisons of continuous variables were performed using two-sided unpaired Student’s *t*-tests or one-way analysis of variance (ANOVA), as appropriate. Correlations between variables were assessed using Pearson’s correlation coefficient. Unless otherwise specified, *p*-values were two-sided and unpaired, and statistical significance was defined as *p* < 0.05. No adjustment for multiple testing was applied for exploratory analyses.

All survival analyses were performed using the survival and survminer packages in R. Data visualization was carried out using the ggplot2 and ComplexHeatmap packages. All statistical analyses were conducted using R (version 4.5.2).

## Results

### Overview of MOSAIC Outputs

The overall inference workflow of the MOSAIC module development framework is summarized in Figure 1. Given a Hematoxylin and Eosin (H&E) whole slide image (WSI) as input, MOSAIC generates tissue segmentation and component-wise, spatially resolved outputs corresponding to the three elements of Nottingham histologic grade (NHG) - mitotic activity, nuclear pleomorphism, and tubule formation, along with an integrated slide-level grade. Detailed descriptions of model development and datasets are provided in the Methods section and Table 1.

**Figure 1:**
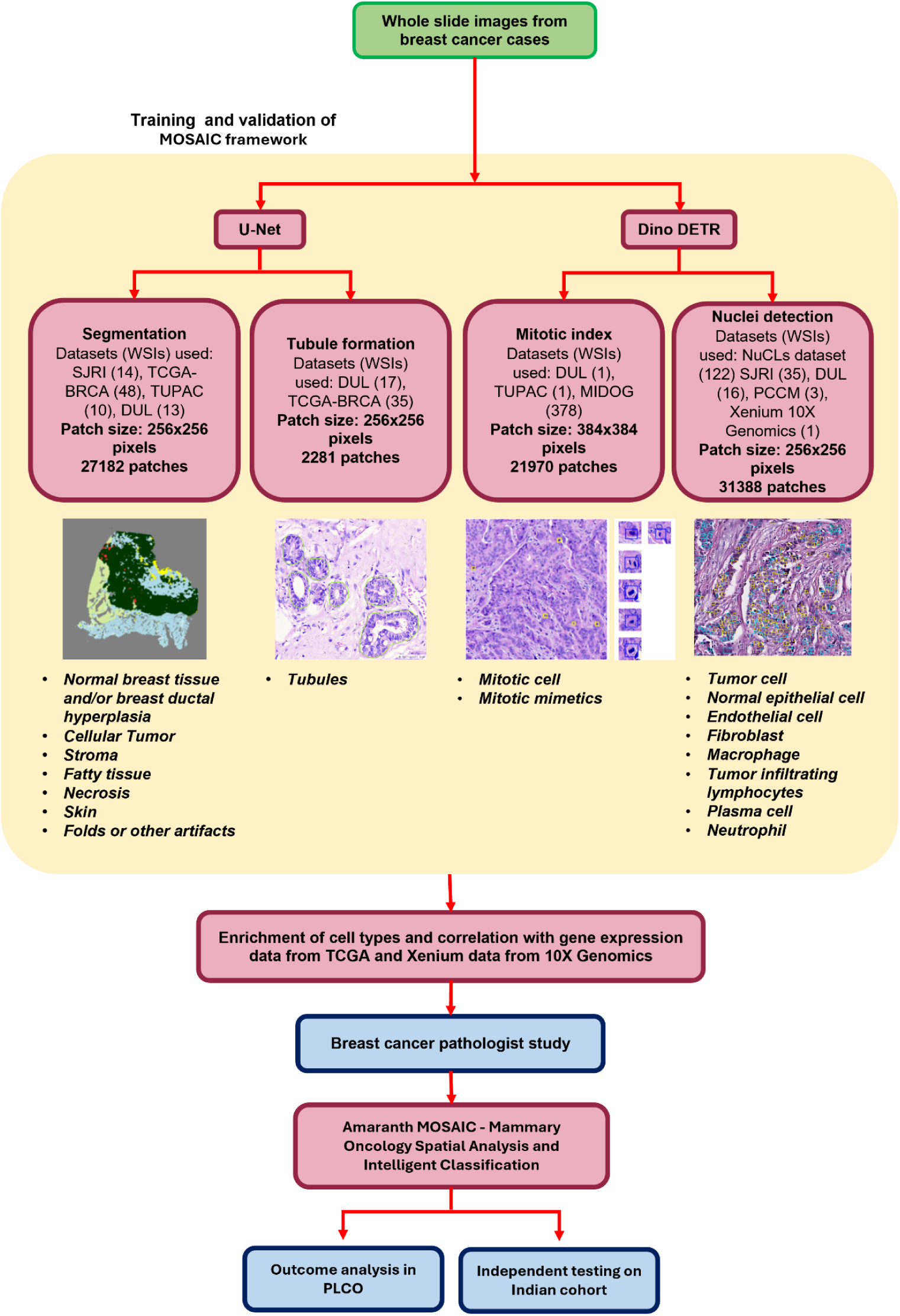
Overview of the MOSAIC module development framework for AI-based Nottingham histologic grading from breast cancer WSIs.

WSIs from breast cancer cases were processed through two deep learning architectures: a U-Net for tissue segmentation and tubule formation detection, and a DINO-DETR model for mitotic activity detection and multi-class nuclei detection.

In the initial stage, MOSAIC performs whole-slide image (WSI) segmentation to delineate distinct anatomical and pathological niches, including cellular tumor (CT), stroma (ST), adipose tissue (FAT), necrosis (NE), skin, and normal breast tissue or ductal hyperplasia (BR-D). The framework quantitatively reports the spatial extent of each segment in pixels, absolute area mm^2^, and as a percentage of the total tissue area, providing a granular architectural profile of the specimen.

For mitotic activity, MOSAIC identifies individual mitotic figures across tumor regions and selects ten consecutive HPFs with the highest mitotic activity. It then provides the spatial overlay of the mitotic cells and HPF regions. Along with spatially localized mitotic detections, the system provides multiple quantitative summaries, including the total mitotic count across the entire WSI, mitotic density expressed as mitoses per mm², mitotic density normalized per 1,000 tumor cells, and the total number of mitotic figures within the selected ten consecutive HPFs. Together, these outputs enable standardized assessment of proliferative activity while preserving spatial context.

For nuclear pleomorphism, MOSAIC detects individual nuclei and assigns cell-type labels like Tumor cells (CE), Normal cells (nCE), Endothelial cells (End), Fibroblasts (fib), Macrophages (Mph), Neutrophils (Neu), Plasma cells (plasma), and Tumor-infiltrating lymphocytes (TIL), enabling cell-type-specific quantification. For each identified cell type, the system reports total cell counts, cell density per mm², and counts normalized per 1,000 tumor cells. In addition, overlap between segmented tissue regions and cell-type detections is used to quantify the proportion of each cell type across histologic compartments. For tumor cells specifically, MOSAIC computes summary statistics of nuclear size, including mean, median, standard deviation, and interquartile range, and generates slide-level histograms depicting the distribution of nuclear sizes across the tumor.

For tubule formation, MOSAIC detects glandular structures across the WSI and contextualizes them based on their spatial relationship to cellular tumor (CT) regions. Quantitative outputs include the total number of tubules within CT regions, the percentage of CT area occupied by tubules, and tubule density expressed as the number of tubules per mm² of tumor area.

In addition to component-level outputs, MOSAIC integrates mitotic activity, nuclear pleomorphism, and tubule formation measurements to generate an overall AI-derived NHG for each case. Importantly, the system retains intermediate quantitative features and spatial overlays for all components, enabling transparent review of the morphologic evidence underlying each component score and the final grade assignment.

Collectively, these outputs provide a structured, interpretable representation of histologic features at both the component and slide levels, forming the basis for subsequent evaluation of technical performance, pathological consistency, and clinical relevance.

### Evaluation of the MOSAIC framework using the TCGA-BRCA cohort

The Cancer Genome Atlas breast cancer cohort (TCGA-BRCA) provides a uniquely comprehensive resource for evaluation of the MOSAIC framework, combining digitized WSIs, pathologist-assigned NHG components, transcriptomic profiles, and clinical outcomes within the same cases. This integration enables simultaneous evaluation of MOSAIC-derived histologic features against expert pathology assessments, underlying molecular signatures, and patient survival.

### Tissue Segmentation

To evaluate MOSAIC-derived tissue segmentation, tissue proportions were compared against independent pathologist annotations and molecular reference measures available in TCGA-BRCA. As illustrated in Figure 2A-B, MOSAIC delineates anatomical compartments directly on digitized H&E WSI, providing spatially resolved overlays alongside quantitative outputs including compartment area (mm²) and percentage of total slide area (Figure 2B-C). Segmentation-derived tissue area percentages demonstrated clear compositional shifts across PAM50 subtypes. Basal-like and HER2-enriched tumors exhibited the highest cellular tumor fractions with comparatively reduced fatty tissue, whereas Luminal A and Normal-like subtypes showed lower tumor proportions and greater stromal and adipose representation. Similar trends were observed across TCGA-BRCA histologic grades, with higher-grade tumors demonstrating increased cellular tumor fraction and relative depletion of non-tumor compartments. These structured compositional differences support the biological validity of the segmentation model in capturing subtype- and grade-associated architectural variation (Supplementary Figure 1A).

**Figure 2:**
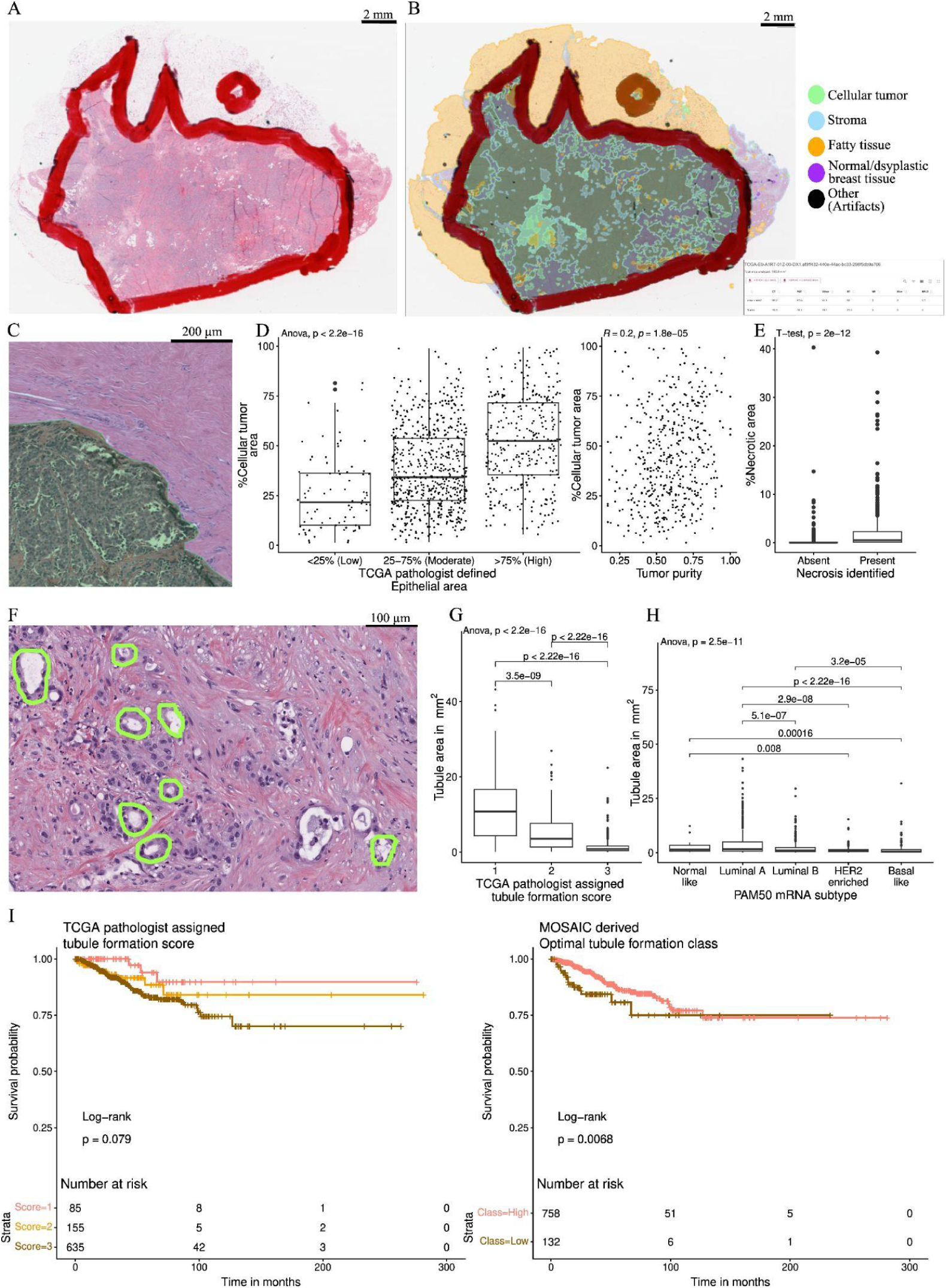
Automated tissue segmentation, quantitative tubule analysis, and survival stratification in TCGA breast cancer. (A) Representative H&E WSI from TCGA-BRCA case TCGA-E9-A1R7-01Z-00-DX1. Scale bar, 2 mm. (B) MOSAIC-derived tissue segmentation overlaid on the WSI, illustrating classification into cellular tumor, stroma, fatty tissue, normal/dysplastic breast tissue, and artifact regions. Quantitative area measurements were computed for each anatomical compartment. Scale bar, 2 mm. (C) High-resolution view demonstrating compartment-level segmentation and discrimination between cellular tumor and stromal regions. Scale bar, 200 µm. (D) Left: Distribution of % CT area across TCGA pathologist-defined epithelial area categories (<25%, 25-75%, >75%). Right: Correlation between % CT area and tumor purity estimated using CIBERSORT. (E) Distribution of % necrotic area stratified by TCGA pathologist annotation of necrosis (absent vs present). (F) Representative H&E WSI from TCGA-A2-A1G6-01Z-00-DX1 showing automated detection and delineation of tubule structures within the cellular tumor compartment. Scale bar, 100 µm. (G) Distribution of tubule area (mm²) across TCGA-BRCA pathologist-assigned tubule formation scores (1-3). (H) Distribution of tubule area (mm²) across PAM50 molecular subtypes (Normal-like, Luminal A, Luminal B, HER2-enriched, Basal-like). (I) Kaplan-Meier survival analysis comparing (left) TCGA-BRCA pathologist-assigned tubule formation scores and (right) optimal binary risk classes derived from the MOSAIC tubule area per mm² metric. Log-rank p-values are shown.

Correlation between automated and manual assessment was evaluated across two histologic features. Cellular tumor fraction increased stepwise across pathologist-defined epithelial content categories (<25%, 25-75%, and >75%), with differences confirmed by one-way ANOVA (p < 2.2 × 10^-16^). Similarly, cellular tumor fraction correlated positively with transcriptome-derived tumor purity estimates from ESTIMATE (R = 0.21, p = 8 × 10^-6^), demonstrating alignment between morphologic and molecular measures of tumor content (Figure 2D). The necrotic area proportion was significantly higher in cases annotated as necrotic by TCGA pathologists compared to those without necrosis (two-sided unpaired t-test, p = 2 × 10^-12^, Figure 2E), confirming that MOSAIC captures tissue-level pathologic features beyond cellular tumor alone.

### Tubule formation

Tubule formation was quantified by MOSAIC through the detection of tubule structures and the calculation of tubule-related features within the cellular tumor compartment (Figure 2F). Across the TCGA-BRCA cohort, multiple tubule-related features were evaluated, including raw tubule counts (using minimum size thresholds of 10,000 pixels), total tubule area (in pixels), percentage of cellular tumor area occupied by tubules, and tubules per mm². All features demonstrated clear and monotonic declines across pathologist-assigned tubule formation scores (one-way ANOVA for all comparisons, *p* < 2.2 × 10^-16^; Supplementary Figure 1B). Among these, tubules per mm² showed the greatest robustness and interpretability across cases and were therefore selected as the defining metric for AI-derived tubule formation scoring (Figure 2G).

Consistent with established biological patterns, tubule density varied across PAM50 molecular subtypes. Tumors classified as normal-like and luminal A demonstrated higher tubule densities, whereas luminal B, HER2-enriched, and basal-like tumors showed progressively lower tubule densities (Figure 2H), reflecting reduced glandular differentiation in more aggressive subtypes.

The prognostic relevance of tubule formation was evaluated in the TCGA-BRCA cohort using both pathologist-assigned and AI-derived classifications. Stratification by pathologist-assigned tubule formation scores showed a trend toward survival separation, although it did not reach statistical significance (log-rank *p* = 0.079; Figure 2I, left). Stratification based on an optimal binary division derived from MOSAIC tubule density per mm^2^ resulted in significant separation of overall survival curves (log-rank *p* = 0.0068; Figure 2I, right), with lower tubule density associated with poorer outcomes.

### Mitotic Activity

Within the TCGA-BRCA cohort, MOSAIC-derived mitotic activity was quantified using five complementary normalizations: mitotic cells across 10 HPFs (0.51 mm diameter each), mitotic cells per 10,000 tumor cells, mitotic cells per mm², total mitotic count per slide, and mitotic cells per 1,000 epithelial cells (Figure 3A). All five measures derived by the MOSAIC mitotic model demonstrated clear stepwise increases across pathologist-assigned mitotic score categories, indicating concordance between AI-derived quantitative measures and manual mitotic grading. One-way ANOVA confirmed significant differences across score groups for each normalization (p < 2.2 × 10^-16^ for all comparisons; Supplementary Figure 2A).

**Figure 3:**
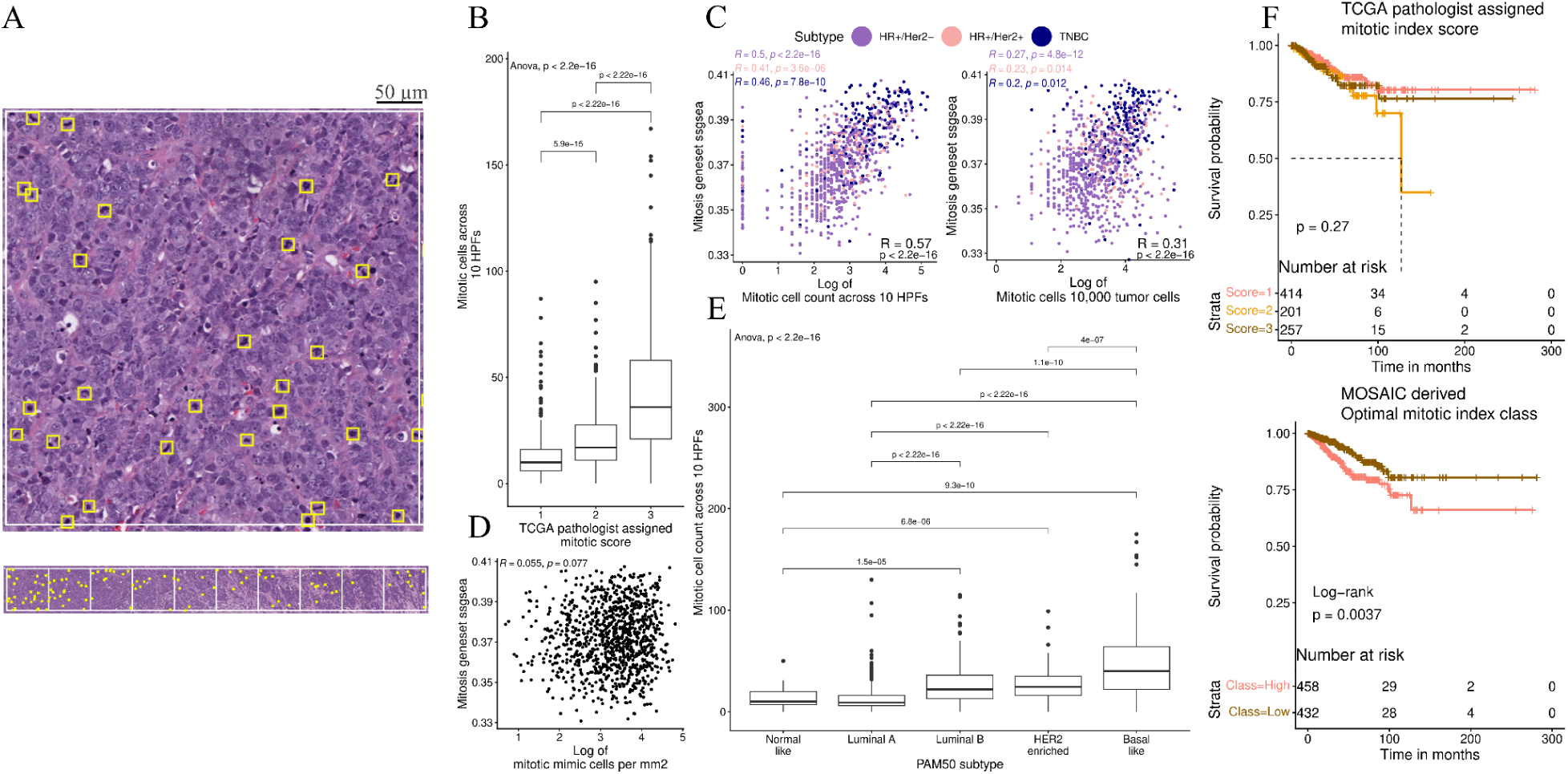
Quantification and clinical relevance of automated mitotic assessment. (A) Representative H&E image from TCGA-BRCA case TCGA-A1-A0SK-01Z-00-DX1 with automated mitotic detections overlaid (yellow boxes). Bottom: spatial distribution of identified HPFs across the corresponding WSI. (B) Distribution of mitotic counts across 10 HPFs stratified by TCGA-BRCA pathologist-assigned mitotic score. (C) Correlation between log-transformed mitotic counts across 10 HPFs (left) and log-transformed mitotic counts per 10,000 tumor cells (right) with ssGSEA enrichment scores derived from the GSEA mitotic gene set. (D) Correlation between log-transformed mitotic mimic density (per mm²) and ssGSEA enrichment scores from the mitotic gene set. (E) Distribution of mitotic counts across 10 HPFs stratified by PAM50 molecular subtype. (F) Kaplan-Meier survival analysis comparing TCGA-BRCA pathologist-assigned mitotic scores (top) and optimal binary risk classes derived from the MOSAIC mitotic count across 10 HPFs (bottom). Log-rank p-values are shown.

Among the evaluated metrics, mitotic cells across 10 HPFs showed the strongest stepwise separation across TCGA-BRCA pathologist-assigned mitotic scores 1, 2, and 3 (Figure 3B). Given the slightly stronger association observed for mitotic cells per 10 HPFs, and its alignment with established clinical scoring practice, this metric was selected as the primary measure for mitotic component scoring.

Accordingly, we assessed its correlation with single-sample gene set enrichment analysis (ssGSEA) scores for a mitosis-related gene set derived from the MSigDB collection. In cases where HPFs were not found, we considered the mitotic cell count per 10,000 tumor cells as both mitotic cells per 10 HPFs and mitotic cells normalized to tumor cell counts showed strong positive correlations with ssGSEA-derived mitosis gene set scores (R = 0.57 and R = 0.31, respectively; both p < 2.2 × 10^-16^, Figure 3C).

During mitotic analysis, MOSAIC explicitly identified a subset of mitotic mimetic nuclei, defined as cells morphologically resembling mitotic figures but lacking characteristic chromatin features of true cell division. In contrast to true mitotic figures, the density of mitotic mimetics showed minimal correlation with mitotic geneset SSGSEA scores (R = 0.055, p-value = 0.077; Figure 3D). This separation highlights the ability of AI-based detection to distinguish proliferative events from non-proliferative morphologic mimics.

MOSAIC-derived mitotic activity also demonstrated subtype-specific trends across PAM50 molecular subtypes. It showed an incremental trend from normal-like and luminal A tumors through luminal B and HER2-enriched tumors, with the highest values observed in basal-like cancers (Figure 3E). This pattern is consistent with known differences in proliferative activity across breast cancer subtypes.

Finally, the prognostic relevance of AI-derived mitotic activity was evaluated in the TCGA-BRCA cohort. Stratification based on pathologist-assigned mitotic scores did not yield significant separation of overall survival curves (log-rank p = 0.27; Figure 3F, top). In contrast, an optimal binary stratification based on MOSAIC-derived mitotic cells per 10 HPFs resulted in significant survival separation (log-rank p = 0.0037; Figure 3F, bottom), with higher AI-derived mitotic activity associated with poorer outcomes.

#### Nuclear pleomorphism

MOSAIC measures nuclear pleomorphism by first identifying and classifying tumor cells. It then measures the size of their nuclei and how much the sizes vary across the tumor.

As part of the pleomorphism module, MOSAIC performs nuclear detection across the tissue sample and classifies them into tumor cells, macrophages, endothelial cells, fibroblasts, lymphocytes, plasma cells, neutrophils, and normal epithelial cells (Figure 4A).

**Figure 4:**
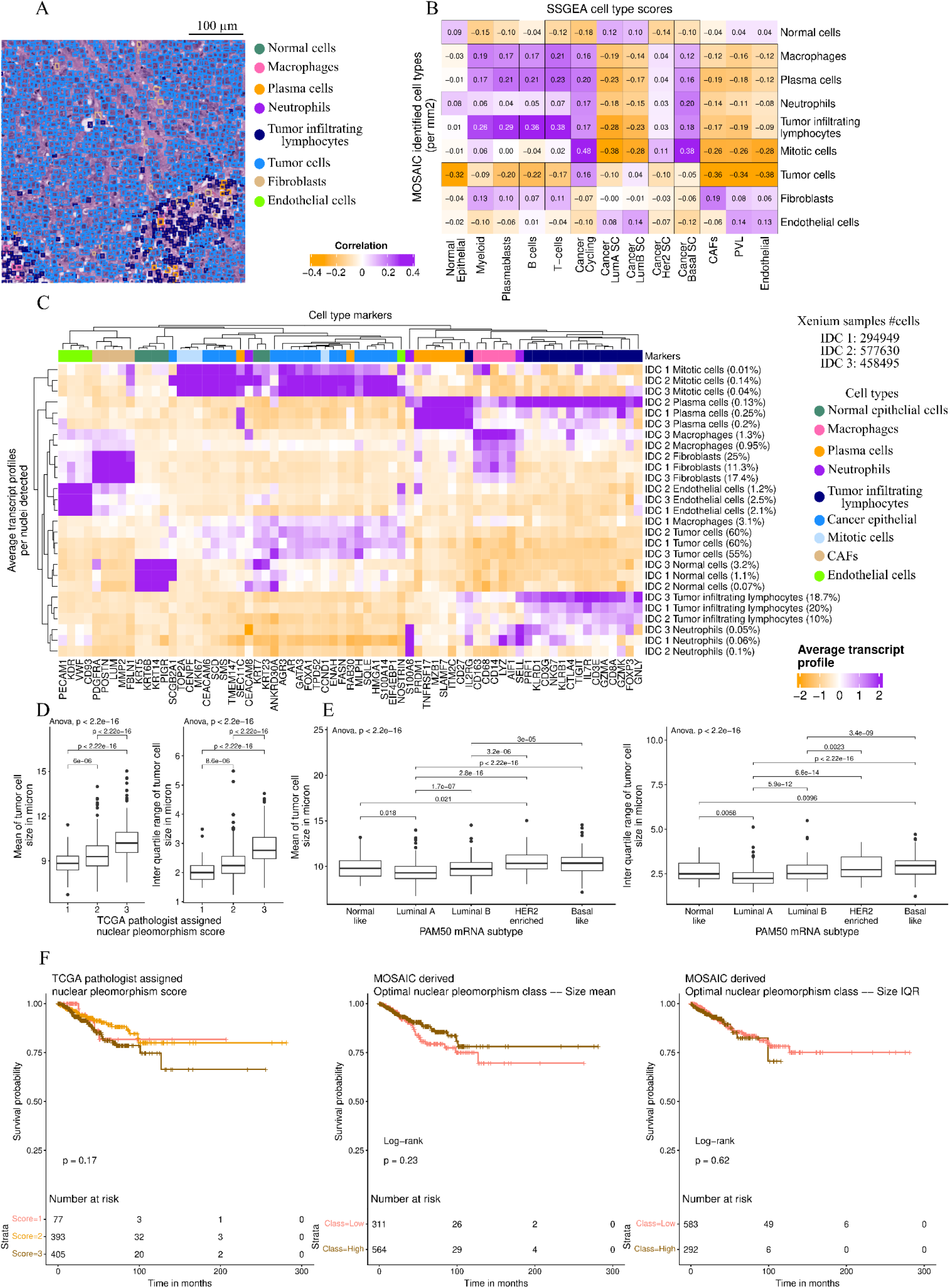
Nuclei detection, cell-type validation, tumor cell morphology, and clinical relevance. (A) Representative H&E image from TCGA-BRCA case TCGA-A1-A0SK-01Z-00-DX1 with MOSAIC nuclei detection overlays. Scale bar, 100 µm. (B) Correlation between MOSAIC-derived cell-type densities (cells per mm²) and ssGSEA enrichment scores of corresponding major cell-type signatures derived from previously published single-cell datasets.^16,17,18^ (C) Heatmap showing average expression of canonical cell-type marker genes and their concordance with MOSAIC-assigned cell types in 10x Xenium spatial transcriptomics data. (D) Distribution of tumor cell size metrics (mean and interquartile range) stratified by TCGA-BRCA pathologist-assigned nuclear pleomorphism score. (E) Distribution of tumor cell size metrics (mean and interquartile range across 10 HPFs) stratified by PAM50 molecular subtype. (F) Kaplan-Meier survival analysis comparing TCGA-BRCA pathologist-assigned nuclear pleomorphism score (left) with optimal binary risk classes derived from MOSAIC tumor cell size mean (middle) and interquartile range (right). Log-rank p-values are shown.

To further validate cell-type assignments, AI-derived cell densities (cells/mm²) were correlated with ssGSEA enrichment scores for major cell-type programs derived from previously published single-cell breast cancer datasets.^16,17,18^ Tumor-infiltrating lymphocyte (TIL) density showed positive correlations with T-cell and B-cell gene signatures (R = 0.38 and 0.36, respectively). Plasmablast density correlated with plasmablast-, T-cell-, and B-cell-associated signatures (R = 0.23, 0.21, and 0.21). Macrophage density was associated with myeloid lineage signatures (R = 0.19), while neutrophil density correlated with basal-like cancer programs (R = 0.20). Fibroblast density showed concordance with cancer-associated fibroblast (CAF) signatures (R = 0.19), and endothelial cell density correlated with endothelial and perivascular lineage programs (R = 0.13 and 0.14). Normal epithelial cell density aligned with normal epithelial and luminal A cancer signatures (R = 0.09 and 0.12), whereas cancer epithelial cell density correlated with cycling cancer programs (R = 0.16). Together, these associations support the biological validity of MOSAIC-derived cell-type quantification (Figure 4B).

To further validate cell-type assignments produced by the MOSAIC nuclei detection model, we applied the trained model to three publicly available breast tissue Xenium images provided by 10x Genomics. These datasets provide spatially resolved transcript counts at single-cell resolution, enabling direct comparison between morphology-based cell-type classification and underlying gene expression profiles.

For each Xenium image, spatial overlap between MOSAIC-detected cell type and Xenium transcripts was used to aggregate transcript counts by predicted cell type, yielding average cell-type-specific transcript expression profiles (Figure 4C).

Distinct and biologically coherent expression patterns were observed across MOSAIC-predicted cell types. Nuclei classified as cancer cells showed enrichment of tumor-associated and proliferative transcripts, including GATA3, TMEM147, and CENPF. Tumor-infiltrating lymphocytes demonstrated elevated expression of immune markers such as FOXP3, IL7R, CD3E, and CD8A, while plasmablast-assigned nuclei were enriched for MZB1 and CD27. Macrophage-assigned nuclei showed increased expression of myeloid-associated genes, including LYZ and CD163, and neutrophil-assigned nuclei exhibited enrichment of S100A8 and SELL. Fibroblast-assigned nuclei were characterized by elevated expression of extracellular matrix-associated transcripts such as POSTN, LUM, and FBLN2, while endothelial-assigned nuclei showed enrichment of vascular markers including VWF, PECAM1, and KDR. Nuclei classified as normal epithelial cells demonstrated enrichment of keratin family transcripts (KRT genes), consistent with non-malignant epithelial identity (Figure 4C). Across the analyzed Xenium images, transcript enrichment was predominantly restricted to the corresponding morphologically defined cell types. Together, these findings provide orthogonal, single-cell resolution validation of MOSAIC’s nuclei classification.

Tumor nuclear morphology was quantified using four continuous measures: mean, median, standard deviation, and interquartile range (IQR) of tumor cell size. All four metrics increased stepwise across TCGA-BRCA pathologist-assigned nuclear pleomorphism scores, with significant differences observed between score categories (one-way ANOVA for all metrics, *p* < 2.2 × 10⁻¹⁶; Supplementary Figure 2B). Mean and median nuclear size showed highly similar distributions across pleomorphism categories, indicating substantial redundancy between these measures (Figure 4D). In contrast, standard deviation exhibited considerable overlap between pleomorphism scores 1 and 2, limiting its discriminative utility. Based on these observations, mean nuclear size and IQR were selected to derive the AI-based pleomorphism score, capturing complementary information on absolute nuclear enlargement and intratumoral variability. Of note, a prominent nucleoli feature was considered, but it showed high inter-pathologist discordance during our pathologist study, and it did not resolve even after repeated discussion amongst pathologists. We decided not to include it in our score calculation.

MOSAIC-derived nuclear morphology also demonstrated clear subtype-specific trends across PAM50 molecular subtypes. Tumor nuclear size differed significantly across subtypes (one-way ANOVA, *p* < 2.2 × 10^-16^), with luminal A tumors exhibiting the smallest median nuclear size, while HER2-enriched and basal-like tumors showed the largest nuclei (Figure 4E). Pairwise comparisons confirmed highly significant differences between luminal A and HER2-enriched (*p* = 2.8 × 10⁻¹⁶) and between luminal A and basal-like tumors (*p* < 2.2 × 10⁻¹⁶), whereas no significant difference was observed between HER2-enriched and basal-like subtypes (*p* = 0.22). These patterns align with established clinicopathologic distinctions, as HER2-enriched and basal-like tumors are typically higher grade and more proliferative, whereas luminal A tumors are generally lower grade. Together, these findings indicate that MOSAIC-derived nuclear features capture biologically meaningful variation in tumor differentiation consistent with intrinsic breast cancer subtypes.

In the TCGA-BRCA cohort, stratification by pathologist-assigned nuclear pleomorphism scores did not yield significant separation of overall survival curves (log-rank *p* = 0.17; Figure 4F, left). Similarly, stratification based on an optimal binary division derived from MOSAIC pleomorphism features did not show significant survival separation (log-rank *p* = 0.23 and p = 0.62; Figure 4F, middle-right), indicating limited prognostic contribution of nuclear pleomorphism alone within this cohort.

### Pathology Calibration and Observer Study

Following internal technical validation and component-wise verification using the TCGA cohort, a multi-phase pathology study was conducted to evaluate the integration of MOSAIC into routine histologic grading workflows and to assess its impact on grading reproducibility. This study was designed to (i) characterize baseline inter- and intra-observer variability in Nottingham grading components, (ii) examine changes in observer concordance with AI-assisted review, and (iii) support calibration of predefined component-level scoring criteria under controlled conditions. Results from this study are presented separately for the controlled Phase 1 evaluation and the large-cohort Phase 2 assessment.

#### Phase 1

Phase 1 was designed as a controlled, repeated-reading study to evaluate intra- and inter-pathologist concordance across sequential grading sessions and to assess how AI assistance influences grading consistency under standardized conditions. A fixed set of 30 breast cancer WSIs was independently reviewed by the same group of seven pathologists across four sessions, including unaided baseline readings (Session 1), repeat unaided readings after washout (Session 2), and two AI-assisted sessions with slide order randomized in all sessions (Session 3 and Session 4). This design enabled separation of baseline variability, intra-observer concordances, and session-wise changes in concordance attributable to AI-supported review.

### Phase 1 intra-pathologist concordance

Intra-pathologist concordance across Phase 1 sessions was evaluated to assess the consistency of individual pathologists when grading the same cases across repeated readings, with and without AI assistance. Weighted Cohen’s κ values were calculated for all pairwise session comparisons for each pathologist and for each Nottingham grading component (Figure 5A). For mitotic activity, baseline intra-observer concordance across unaided sessions (Session 1 vs Session 2) showed fair to moderate repeatability, with weighted κ values ranging from 0.07 to 0.59. Following the introduction of MOSAIC assistance, intra-observer concordance improved consistently across all readers, with AI-assisted session comparisons (Session 3 vs Session 4) yielding κ values between 0.66 and 0.78, indicating more stable mitotic scoring across repeated evaluations. Nuclear pleomorphism demonstrated comparatively higher baseline repeatability, with unaided κ values ranging from 0.33 to 0.71, corresponding to moderate to substantial agreement. AI assistance did not result in a systematic change for this component, as AI-assisted κ values largely overlapped with baseline ranges (0.33 - 0.69). In contrast, tubule formation exhibited the greatest baseline intra-observer variability, with κ values ranging from 0.28 to 0.80 across unaided session comparisons. With AI assistance, tubule formation scoring became more consistent, with κ values increasing to 0.35-0.91 in AI-assisted sessions, and several readers achieving substantial to near-perfect repeatability. Overall, Phase 1 intra-observer analyses indicate that AI assistance substantially improved the repeatability of mitotic activity and tubule formation scoring, particularly among readers with lower baseline consistency, while nuclear pleomorphism showed relatively stable repeatability irrespective of AI support (Figure 5A).

**Figure 5:**
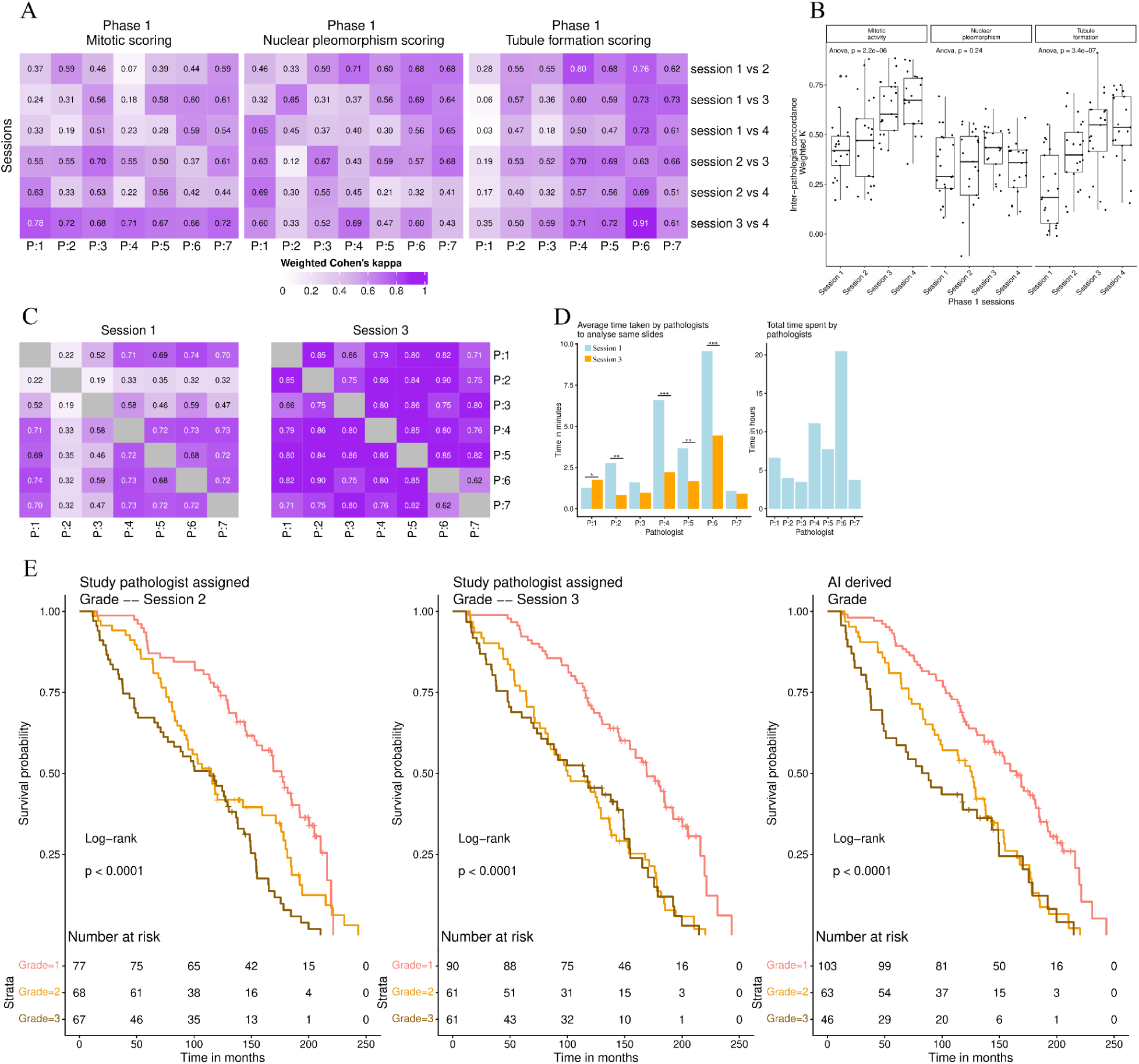
Pathologist concordance, grading efficiency, and survival validation with AI assistance. **(A)** Intra-pathologist concordance (n = 7) across four sessions in Phase 1, evaluated using weighted Cohen’s kappa for the three grading components: mitotic activity, nuclear pleomorphism, and tubule formation. **(B)** Inter-pathologist concordance (n = 7) across four sessions in Phase 1, evaluated using weighted Cohen’s kappa for mitotic activity, nuclear pleomorphism, and tubule formation. **(C)** Inter-pathologist concordance for overall grade in Phase 2, comparing Session 1 (unassisted) and Session 3 (AI-assisted), evaluated using weighted Cohen’s kappa. **(D)** Time required by each pathologist to complete grading in Session 1 versus Session 3, demonstrating changes in grading efficiency with AI assistance. **(E)** Kaplan-Meier survival analysis of 212 patients from the NIH-PLCO breast cancer cohort stratified by three-grade categories: Session 1 pathologist-assigned grades (left), Session 3 AI-assisted pathologist-assigned grades (middle), and fully AI-derived grades (right).

### Phase 1 inter-pathologist concordance

Inter-pathologist concordance in Phase 1 was assessed by calculating weighted Cohen’s κ across all reader pairs for each Nottingham grading component and comparing distributions across sequential sessions (Figure 5B). For mitotic activity, inter-observer agreement increased progressively across sessions, with median κ values shifting upward from Session 1 through Session 4, indicating a consistent improvement in concordance with repeated evaluation and AI assistance. This increase was statistically significant at the group level (ANOVA test, *p* = 2.2 × 10⁻⁶).

A similar pattern was observed for tubule formation, where inter-pathologist concordance showed a marked upward shift across sessions, with higher median κ values and reduced dispersion in later sessions, also reaching statistical significance (ANOVA test, *p* = 3.4 × 10⁻^7^). In contrast, nuclear pleomorphism demonstrated relatively stable inter-observer agreement across sessions, with overlapping κ distributions and no significant session-wise change (ANOVA test, *p* = 0.24). Collectively, these results indicate that inter-pathologist agreement improved across Phase 1 sessions for mitotic activity and tubule formation, while nuclear pleomorphism remained comparatively unchanged (Figure 5B).

### Derivation of Component-Level Scoring Thresholds

Data from the Phase 1 sessions, conducted with AI assistance, were used to examine the relationship between MOSAIC-derived quantitative features and pathologist-assigned scores for mitotic activity, nuclear pleomorphism, and tubule formation. Distributions of continuous AI features were evaluated across manual score categories to identify consistent quantitative ranges and inform the definition of component-level scoring thresholds. Thresholds were finalized after Phase 1 and were provided as AI assistance in Phase 2 session 3.

### Phase 2

Phase 2 was designed to evaluate the scalability and generalizability of AI-assisted grading in a larger, clinically representative cohort. The NIH-PLCO (Prostate, Lung, Colorectal, and Ovarian Cancer Screening Trial) dataset comes from a large U.S. randomized study that followed ∼155,000 adults to evaluate cancer screening methods and long-term health outcomes, including cancer incidence and mortality. Their breast cancer dataset consists of participants who developed breast cancer during the course of the study. Using WSIs from the NIH-PLCO breast cancer cohort, the same group of seven pathologists participated in three grading sessions. In Session 1, all pathologists independently graded a common set of 41 images without AI assistance to establish baseline inter-observer variability. In Session 2, each pathologist graded a unique set of 30 images, allowing assessment of individual grading behavior and intra-observer reference without AI support. Session 3 consisted of 212 images, forming a superset that included all images from Session 2, and was evaluated with AI assistance enabled.

### Phase 2 inter-pathologist concordance

Inter-pathologist concordance in Phase 2 was assessed by comparing weighted Cohen’s κ values between baseline unaided grading (Session 1) and AI-assisted grading at scale (Session 3) across all Nottingham grading components and the overall grade. For mitotic activity, inter-observer agreement increased substantially, with κ values ranging from 0.18-0.70 in Session 1 and shifting to 0.79-1.00 in Session 3, indicating a transition from fair-moderate to substantial-near-perfect agreement (Supplementary Figure 3A). Tubule formation showed a similar improvement, with κ values increasing from 0.03-0.81 at baseline to 0.58-0.88 with AI assistance (Supplementary Figure 3B). Nuclear pleomorphism demonstrated comparatively stable but narrower gains, with κ values ranging from 0.03-0.77 in Session 1 and 0.47 -0.79 in Session 3 (Supplementary Figure 3C). When integrating all components into an overall AI-derived grade, inter-pathologist concordance improved markedly, with κ values increasing from 0.19-0.74 in Session 1 to 0.62-0.90 in Session 3 (Figure 5C). Collectively, these findings demonstrate that AI assistance leads to consistent and substantial improvements in inter-pathologist agreement for mitotic activity, tubule formation, and overall grading when applied at scale, while nuclear pleomorphism remains comparatively less responsive to AI-driven standardization.

### AI Assistance Reduced Cognitive Load and Turnaround Time

During Phase 2, pathologists frequently reported increased difficulty when evaluating slides characterized by suboptimal staining, limited tumor content, poor tissue preservation, or ambiguous invasive components. Common challenges included distinguishing degenerating or apoptotic cells from true mitotic figures, interpreting nuclear morphology in under- or over-stained regions, and assessing tubule formation in fragmented or sparsely cellular tissue. In these scenarios, AI assistance reduced cognitive load by providing stable, consistent visual annotations and prioritizing regions of interest, minimizing the need for exhaustive manual scanning across large tissue areas.

Consistent with these qualitative observations, AI assistance was associated with a reduction in grading time for most participants (Figure 5D). Average per-slide review time decreased in Phase 2 relative to baseline unaided sessions, with the largest reductions observed among readers who initially required longer evaluation times. For example, one pathologist (P6) reduced mean grading time from approximately 9 minutes per slide in Session 1 to approximately 4 minutes per slide in the AI-assisted session. While some observers (e.g., P3 and P7) demonstrated minimal change in review time, the overall trend across participants indicated improved workflow efficiency with AI support, suggesting reduced mental fatigue and faster convergence on grading decisions, particularly in challenging cases.

### Prognostic Impact of AI-Assisted and AI-Derived Grading

Across all three grading approaches, pathologist-assigned (Session 2), AI-assisted pathologist grading (Session 3), and fully AI-derived grading, histologic grade demonstrated significant stratification of patient survival in the NIH-PLCO breast cancer cohort(log-rank *p* < 0.0001 for all comparisons; Figure 5E).

Comparative inspection of the survival curves revealed differences in how patients were distributed across risk groups. In the pathologist-only session, survival curves for Grades 2 and 3 showed partial overlap during mid follow-up, indicating limited separation between intermediate- and high-risk disease. With AI-assisted grading, the separation between Grade 1 and higher grades became more pronounced, while maintaining ordinal separation across all three grades. In the fully AI-derived grading, Grade 1 demonstrated early and sustained separation from Grades 2 and 3, and the Grade 3 group formed a smaller, more distinct high-risk cohort.

These shifts were reflected in patient distribution across grades. Relative to pathologist-only grading, AI-derived grading increased the number of cases assigned to Grade 1 (77 to 103) and reduced the number classified as Grade 3 (67 to 46). Importantly, the survival trajectories of these reclassified groups aligned with their assigned grades, suggesting that AI-driven refinement of grading thresholds improves the clarity of prognostic stratification, particularly at the low-intermediate and intermediate-high grade boundaries (Figure 5E).

Together, the Phase 1 and Phase 2 pathology studies demonstrate that MOSAIC-assisted grading improves intra- and inter-pathologist concordance for mitotic activity and tubule formation, stabilizes grading across repeated evaluations, and reduces review time in a large, heterogeneous cohort. Phase 1 provided the basis for defining component-level scoring thresholds, which were subsequently applied consistently during Phase 2 evaluations.

Building on these observer-based findings, we next evaluated the standalone application of AI-derived component scores and overall grades to entire independent cohorts. This analysis was designed to assess grading concordance and prognostic performance when MOSAIC is applied uniformly, without pathologist interaction, across diverse datasets.

### Application of AI-Derived Grades Across Independent Cohorts

Following threshold finalization, MOSAIC-derived component scores and overall grades were applied to entire breast cancer cohorts from TCGA-BRCA, NIH-PLCO breast cancer cohort, and St. John’s Research Institute (SJRI). These cohorts were selected to represent complementary validation contexts, including a public dataset with comprehensive clinical and pathology annotation (TCGA), a large population-based cohort with long-term survival follow-up (NIH-PLCO), and an institutional Indian cohort (SJRI).

For each dataset, MOSAIC was applied to WSIs to generate component-level scores for mitotic activity, nuclear pleomorphism, and tubule formation, as well as an integrated AI-derived NHG. In the TCGA-BRCA cohort, AI-derived component scores and overall grades were evaluated against pathologist-assigned component scores and overall grade, together with survival outcomes. In the NIH-PLCO breast cancer and SJRI cohorts, analyses focused on concordance with pathologist-assigned overall grade and on survival stratification, as component-level pathologist scores were not available. All analyses used the same scoring criteria defined after Phase 1, without dataset-specific recalibration. All the parameters derived by MOSAIC in individual cohorts are provided in Supplementary File 2.

### TCGA-BRCA cohort: Concordance of MOSAIC-Derived Grading With TCGA Pathologist Assessment

Concordance between MOSAIC-derived scores and TCGA-BRCA pathologist-assigned assessments varied across individual NHG components but was strongest for tubule formation and the integrated overall grade. Tubule formation demonstrated the highest agreement (accuracy = 0.6607; Cohen’s κ = 0.549), with a pronounced diagonal pattern in the confusion matrix, particularly for Score 3, indicating robust identification of poorly differentiated architecture. Mitotic activity showed moderate concordance (accuracy = 0.4985; κ = 0.40), with most disagreement arising between Scores 1 and 2, consistent with known challenges in distinguishing low-range proliferative activity. Nuclear pleomorphism exhibited the lowest agreement (accuracy = 0.3303; κ = 0.271), characterized by substantial dispersion across score categories, especially within intermediate-grade cases, underscoring the intrinsic subjectivity and reduced reproducibility of this feature. Despite variability at the component level, the integrated AI-derived overall grade remained comparatively stable (accuracy = 0.5637; κ = 0.539) (Figure 6A).

**Figure 6:**
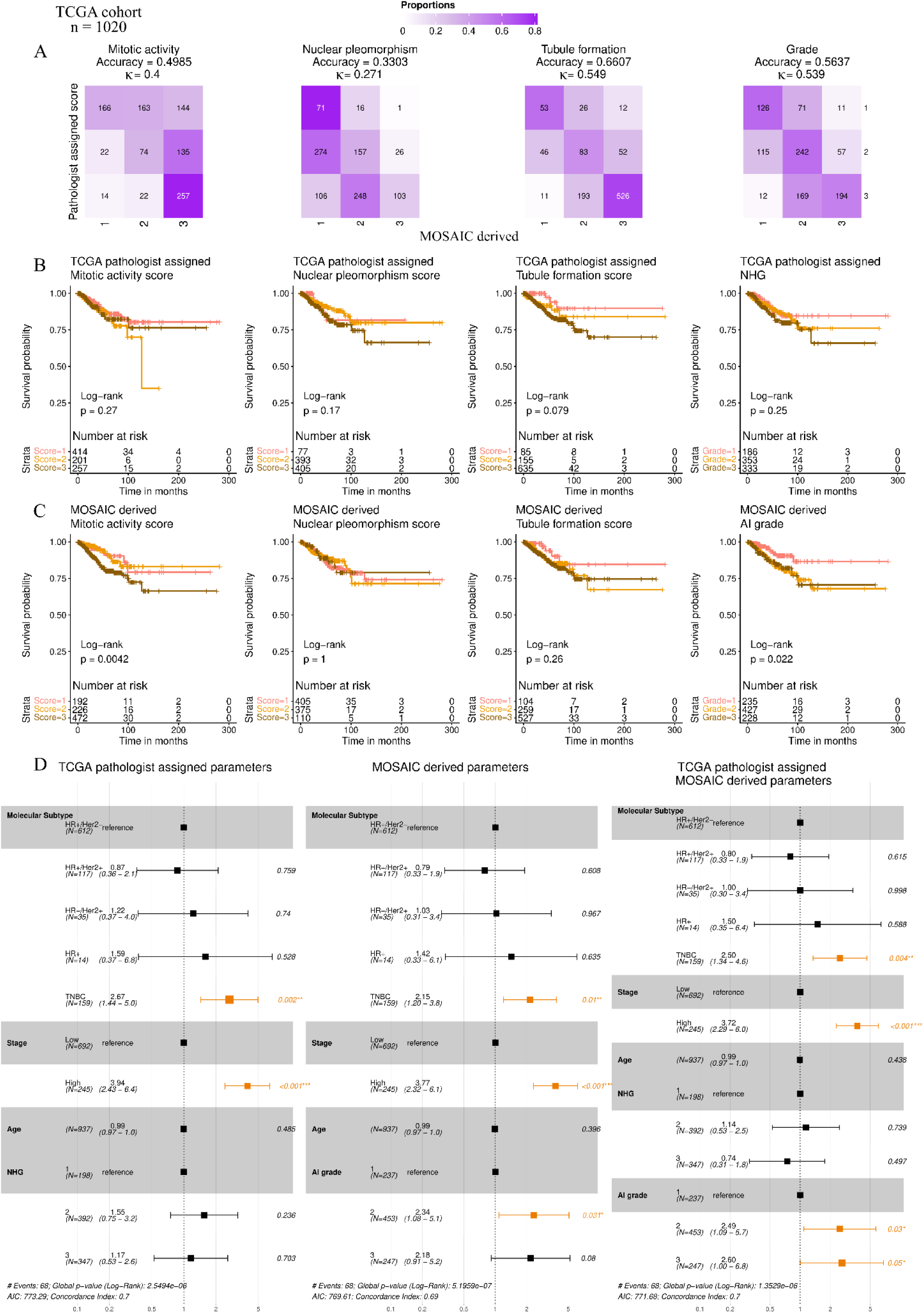
Concordance, prognostic stratification, and multivariable survival analysis of MOSAIC-derived grading in the TCGA-BRCA cohort (n = 1,020). (A) Confusion matrices comparing TCGA-BRCA pathologist-assigned scores for mitotic activity, nuclear pleomorphism, and tubule formation, as well as overall histologic grade, with corresponding MOSAIC-derived scores. Concordance was quantified using weighted Cohen’s kappa, and accuracy was calculated using pathologist-assigned scores as the reference. (B) Kaplan-Meier survival analysis stratified by pathologist-assigned mitotic activity, nuclear pleomorphism, tubule formation scores, and overall grade. (C) Kaplan-Meier survival analysis stratified by MOSAIC-derived mitotic activity, nuclear pleomorphism, tubule formation scores, and overall grade. (D) Multivariable Cox proportional hazards analysis incorporating clinical covariates, including age, stage, and molecular subtype. Forest plots show models using pathologist-assigned grade (left), MOSAIC-derived grade (middle), and a combined model including both grades (right).

In the TCGA-BRCA cohort, the prognostic relevance of MOSAIC-derived grading was evaluated using multiple survival endpoints, including overall survival (OS), disease-free interval (DFI), disease-specific survival (DSS), and progression-free interval (PFI). AI-derived histologic grade and individual component scores showed consistent trends of risk stratification across endpoints. For subsequent analyses, DFI was used as the primary survival endpoint, as it demonstrated the most consistent separation across grades from Grade 1 through Grade 3 (Supplementary File 3).

In the TCGA-BRCA cohort, pathologist-assigned NHG did not significantly stratify DFI, whereas the MOSAIC-derived overall grade showed a statistically significant separation of outcomes (*p* = 0.022, Figure 6B-C). A similar pattern was observed at the component level for mitotic activity, where the pathologist-assigned mitotic score was not associated with DFI, while the AI-derived mitotic score showed a significant prognostic association (*p* = 0.0042, Figure 6B-C). In contrast, nuclear pleomorphism and tubule formation did not achieve statistical significance for survival stratification using either pathologist assigned or AI-derived scoring.

In the TCGA HR+/HER2− cohort, survival and Cox proportional hazards analyses were performed using both TCGA-BRCA pathologist-assigned component scores and overall grade, as well as MOSAIC-derived scores and grade. Pathologist-assigned scores and grades showed substantial overlap in survival curves across the three strata, limiting prognostic separation. In contrast, MOSAIC-derived metrics demonstrated stronger stratification trends, with nuclear pleomorphism (p = 0.065) and tubule formation (p = 0.054) approaching statistical significance and showing visible separation of survival curves, particularly identifying individuals with score 3 and poorer outcomes. Similarly, MOSAIC-derived overall grade showed near-significant prognostic separation (p = 0.066), with clearer survival differences for Grade 1 cases relative to higher grades (Supplementary Figure 4A-B).

Multivariable Cox regression analyses comparing pathologist-assigned grading, AI-derived grading, and their combination showed that AI-assisted pathology grading provided the most informative prognostic model. In models including only pathologist-assigned NHG, grade was not independently associated with survival (Figure 6D - left). In contrast, models incorporating AI-derived grade demonstrated a significant association with outcome (Figure 6D - middle).

When both pathologist-assigned NHG and AI-derived grade were included in the same model, AI grade retained independent prognostic significance, whereas NHG remained non-significant (Figure 6D - right). This suggests that AI-derived grading captures prognostically relevant morphologic information that is not fully represented in conventional manual grading. Model-level performance metrics further supported this observation: the AI-derived and AI-assisted models showed improved global likelihood statistics and lower Akaike Information Criterion (AIC) values compared with the pathologist-only model, while maintaining comparable concordance indices. Collectively, these findings indicate that AI-assisted grading augments traditional pathology by refining risk stratification without displacing established clinical predictors such as stage, supporting its role as a complementary decision-support tool rather than a replacement for expert assessment. Similar trends were observed in the TCGA-BRCA HR+/Her2-sub-population (Supplementary Figure 4C).

#### NIH-PLCO Breast Cancer Cohort: Concordance and Survival Stratification Using AI-Derived Grading

The NIH-PLCO breast cancer cohort was used to evaluate the performance of MOSAIC-derived grading in a large, population-based setting with long-term survival follow-up. Unlike TCGA-BRCA, component-level pathologist scores were not available; therefore, analyses focused on concordance between MOSAIC-derived and pathologist-assigned overall NHG and on the prognostic stratification achieved by both. Kaplan-Meier survival analyses were performed using MOSAIC-derived scores for mitotic activity, nuclear pleomorphism, and tubule formation to assess their independent associations with outcome.

Concordance analysis demonstrated a clear diagonal pattern, indicating substantial agreement across grade categories. Agreement was strongest for Grade 1 and Grade 3 tumors, with most discordance occurring between adjacent grades (κ = 0.539, Figure 7A). Importantly, extreme misclassifications were uncommon, suggesting that MOSAIC largely preserves the ordinal structure of NHG while introducing limited shifts near clinical decision boundaries rather than at the extremes.

**Figure 7:**
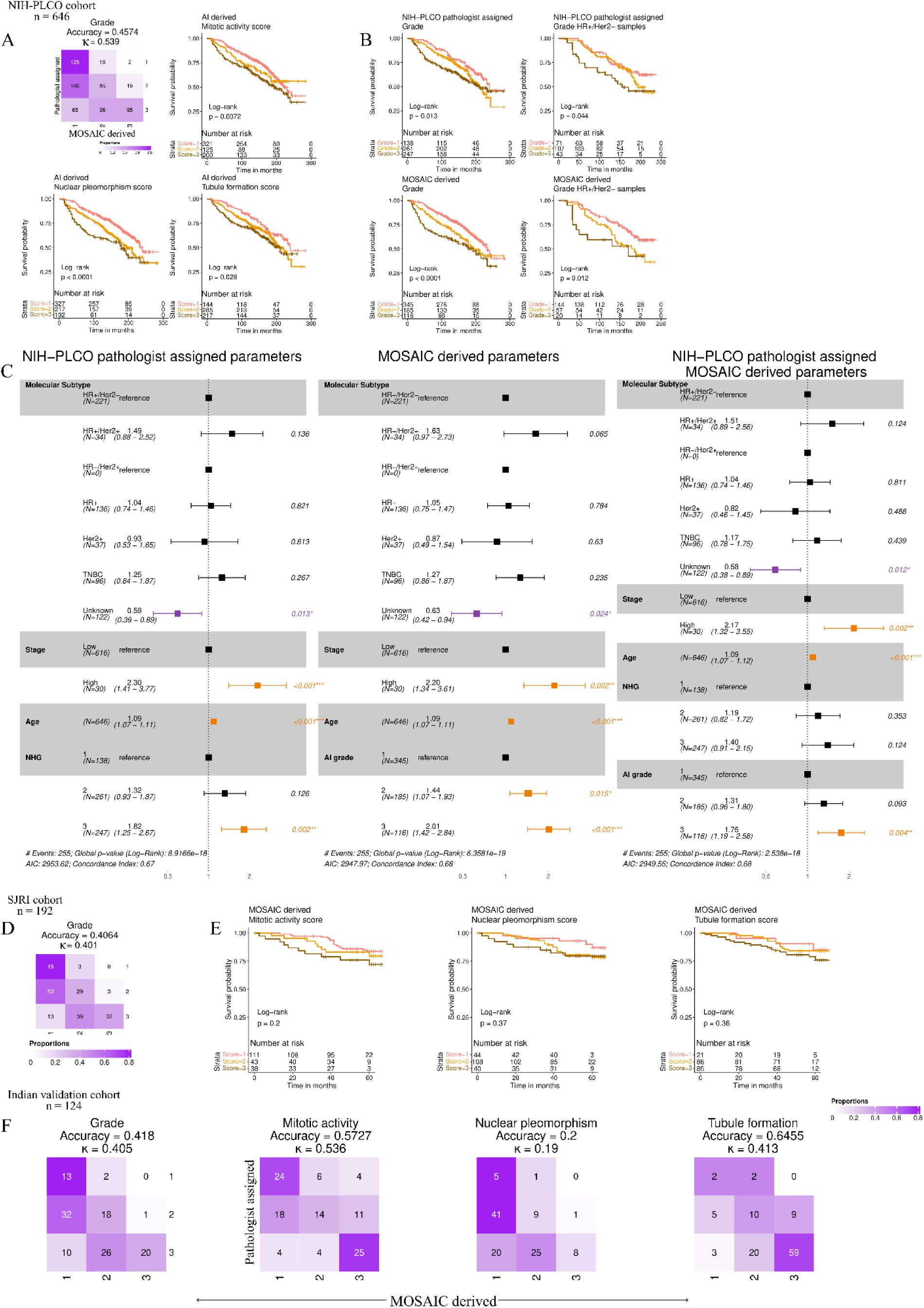
External and internal validation of MOSAIC-derived grading across NIH-PLCO, SJRI, and multi-institutional cohorts. This figure includes survival and concordance analyses across three independent cohorts: the NIH-PLCO breast cancer cohort (n = 646), the SJRI cohort (n = 192), and an internal multi-institutional validation cohort (n = 125) comprising cases from Samved, Zydus, UL, and BVDUMC. (A) Concordance between NIH-PLCO pathologist-assigned grades and MOSAIC-derived grades shown as a confusion matrix. Kaplan-Meier survival curves stratified using MOSAIC-derived mitotic activity, nuclear pleomorphism, and tubule formation scores are shown alongside. (B) Kaplan-Meier survival analysis in the NIH-PLCO breast cancer cohort. The top row shows survival stratified by NIH-PLCO pathologist-assigned grades in the complete cohort and in the HR+/HER2− subset. The bottom row shows survival stratified by MOSAIC-derived grades for the same groups. (C) Multivariable Cox proportional hazards analysis including clinical covariates (age, stage, and molecular subtype). Forest plots show models using NIH-PLCO pathologist-assigned grade (left), MOSAIC-derived grade (middle), and a combined model including both pathologist and MOSAIC grades (right). (D) Validation in the SJRI cohort (n = 192), showing concordance between pathologist-assigned and MOSAIC-derived grades and Kaplan-Meier survival curves using MOSAIC-derived mitotic activity, nuclear pleomorphism, and tubule formation scores. (E) Concordance between pathologist-assigned scores and grades and MOSAIC-derived scores and grade in the internal multi-institutional validation cohort (n = 124).

Survival analyses showed that all three MOSAIC-derived grading components were significantly associated with patient outcome (Figure 7B). Direct comparison of survival stratification using NIH-PLCO pathologist-assigned versus MOSAIC-derived overall grades further highlighted the added prognostic resolution provided by AI-based grading. In the larger NIH-PLCO cohort, pathologist-assigned grading showed a statistically significant association with survival (log-rank p = 0.013), but survival curves for Grades 1 and 2 remained closely spaced, indicating limited discrimination between low- and intermediate-risk disease. In contrast, MOSAIC-derived grading achieved markedly stronger separation (log-rank p < 0.0001), driven by a clear and early divergence of Grade 1 tumors from higher grades and improved ordering across all three risk groups (Figure 7B).

This advantage was more pronounced within the HR+/HER2- subtype. Pathologist-assigned grading reached statistical significance (log-rank p = 0.044) but showed substantial overlap between Grades 1 and 2. By comparison, MOSAIC-derived grading provided clearer separation of Grade 1 tumors (log-rank p = 0.012), with reduced overlap and more consistent stratification of low-risk disease (Figure 7B). Collectively, these results indicate that MOSAIC-derived grading improves prognostic resolution beyond manual assessment in the NIH-PLCO breast cancer cohort, particularly in clinically challenging settings where conventional grading struggles to distinguish adjacent risk categories.

Multivariable Cox proportional hazards models were constructed to compare the independent prognostic contributions of NIH-PLCO pathologist-assigned NHG and MOSAIC-derived AI grade after adjustment for age, stage, and molecular subtype. In models including clinical variables and NIH-PLCO pathologist-assigned NHG, neither Grade 2 nor Grade 3 showed a significant association with survival (Grade 2: HR 1.31, 95% CI 0.93-1.86, p = 0.124; Grade 3: HR 1.35, 95% CI 0.89-2.07, p = 0.158) (Figure 7C). In contrast, when NHG was replaced with the AI-derived grade, AI Grade 2 emerged as an independent predictor of risk (HR 1.39, 95% CI 1.04-1.86, p = 0.024), while higher stage and increasing age remained significant covariates. In a combined model incorporating both pathologist-assigned NHG and AI-derived grade, the AI grade retained prognostic relevance, whereas pathologist NHG remained non-significant, indicating that MOSAIC-derived grading captures prognostic information beyond that provided by conventional Nottingham histologic grading (Figure 7C).

#### MOSAIC Validation in independent Indian cohorts

The SJRI cohort was used to evaluate the performance of MOSAIC-derived grading in an institutional Indian setting with available clinical follow-up. MOSAIC was applied to surgical WSIs to generate component-level scores for mitotic activity, nuclear pleomorphism, and tubule formation, as well as an integrated AI-derived NHG, using the same scoring criteria established after Phase 1.

Concordance analysis in the SJRI cohort demonstrated moderate agreement between MOSAIC-derived and pathologist-assigned overall NHG, with most discrepancies occurring between adjacent grade categories (Figure 7D). This pattern reflects the known biological and interpretive heterogeneity of intermediate-grade tumors and mirrors observations from TCGA-BRCA and NIH-PLCO breast cancer cohorts.

Overall, MOSAIC preserved the ordinal structure of Nottingham grading in the SJRI cohort, with disagreements largely confined to clinically adjacent grades rather than reflecting systematic over- or under-grading. Survival analyses were exploratory in the SJRI cohort, and even though they did not demonstrate statistically significant separation across grades, the trend was similar to TCGA and NIH-PLCO data. This is attributable to the limited sample size, low event rate, and the fact that median survival was not reached in any grade group during the available follow-up (Figure 7E). Although survival analyses in the SJRI cohort did not reach statistical significance, MOSAIC-derived grading showed clearer visual separation of Grade 3 tumors from lower grades, whereas survival curves based on pathologist-assigned NHG demonstrated substantial overlap across all three grade categories (Supplementary Figure 5A-B).

In the SJRI cohort, multivariable Cox regression analyses demonstrated that MOSAIC-derived grade showed a statistically significant association with survival when evaluated alongside clinical covariates, with Grade 2 tumors exhibiting increased risk relative to Grade 1 (HR ≈ 1.39, *p* = 0.02). In contrast, pathologist-assigned NHG did not show a significant independent association with outcome when modeled alone. When both MOSAIC-derived grade and pathologist-assigned grade were included in the same model, neither grading variable retained statistical significance, while clinical stage remained the dominant predictor of outcome (HR ≈ 4.14, *p* < 0.001). Molecular subtype and age were not independently associated with survival in any model. These findings suggest that MOSAIC-derived grading captures prognostic information in this cohort, but its effect overlaps with conventional grading when jointly modeled, likely reflecting shared variance and limited statistical power due to small sample size and low event rates (Supplementary Figure 5C).

Concordance analysis was also conducted on a combined Indian cohort comprising cases from Dr. Udayan Laboratory (DUL), Bharati Vidyapeeth Deemed University Medical College (BVDUMC), Samved Medicare Hospital (SMH), and Zydus Hospitals (ZH). Agreement between MOSAIC-derived scores and pathologist-assigned Nottingham grading ranged from fair to moderate across individual components. Tubule formation showed the highest concordance (accuracy = 0.6607; Cohen’s κ = 0.544), followed by overall grade (accuracy = 0.5637; κ = 0.539), both corresponding to moderate agreement.

Mitotic activity demonstrated moderate agreement (accuracy = 0.4985; κ = 0.40), while nuclear pleomorphism showed fair agreement (accuracy = 0.3303; κ = 0.271). Across all parameters, most discordance occurred between adjacent grades, indicating preservation of ordinal grading structure in Indian settings characterized by heterogeneous staining processes and scanning platforms (Figure 7F).

## Discussion

In this study, we introduce MOSAIC, an explainable AI framework designed to improve the reproducibility and prognostic utility of Nottingham histologic grading in breast cancer. Prior AI-based grading studies have demonstrated that deep learning models can approximate or match pathologist grading performance and show prognostic relevance when applied retrospectively to curated datasets.^19,20^ MOSAIC extends this body of work by standardizing component-level assessment of mitotic activity, nuclear pleomorphism, and tubule formation from WSIs, while preserving the established Nottingham grading paradigm and explicitly evaluating AI integration into pathologist workflows.

A key finding of this work is that the impact of AI assistance is component dependent, consistent with prior reports of differential reproducibility across Nottingham grading components.^1,21^ Mitotic activity and tubule formation showed the greatest gains in reproducibility and prognostic relevance, whereas nuclear pleomorphism remained comparatively resistant to standardization. This divergence likely reflects intrinsic differences in how these features are assessed. Mitotic activity and tubule formation rely on spatial sampling, field selection, and threshold-based counting, which are particularly susceptible to observer fatigue and search bias, and which AI systems are well-suited to standardize.^8,10^ In contrast, nuclear pleomorphism remains inherently subjective, dominated by intermediate categories and lacking universally accepted quantitative thresholds, a limitation consistently noted in both manual grading studies and AI-based approaches.^19,20^ The modest gains observed for pleomorphism, therefore, likely reflect fundamental limitations of the criterion rather than model performance.

The multi-phase observer study provides direct evidence for the role of MOSAIC in reducing grading variability under realistic diagnostic conditions. Unlike prior AI grading studies that primarily evaluate stand-alone model performance,^5,19^ MOSAIC explicitly assesses AI-assisted grading. In Phase 1, AI assistance improved intra- and inter-observer concordance for mitotic activity and tubule formation across repeated readings, with the largest gains observed among readers with lower baseline consistency. Phase 2 demonstrated that these improvements were preserved at scale in a larger, heterogeneous cohort, while also reducing average grading time for many participants. These findings align with emerging evidence that AI can reduce cognitive load and search bias in digital pathology without altering diagnostic responsibility.^20^

Beyond reproducibility, MOSAIC-derived grading demonstrated meaningful prognostic relevance. In the TCGA-BRCA cohort, AI-derived overall grade and mitotic score stratified disease-free interval more effectively than pathologist-assigned NHG, and multivariable Cox regression showed that AI-derived grade retained independent prognostic significance after adjustment for stage and molecular subtype.

Prior studies have reported prognostic equivalence between AI-based grading and manual grading,^5,19^ but MOSAIC further demonstrates that quantitative component integration can yield prognostic information not captured by conventional grading alone, particularly at clinically ambiguous grade boundaries.

The generalizability of MOSAIC was supported by consistent performance across diverse datasets. TCGA-BRCA provided a richly annotated reference cohort, NIH-PLCO breast cancer cohort enabled validation in a large population-based setting with long-term follow-up, and the combined Indian cohort represented heterogeneous real-world diagnostic environments with variability in staining, scanning platforms, and institutional practice. Many prior AI grading studies rely on single-cohort or single-institution validation.^19,20^ By contrast, MOSAIC demonstrated preserved ordinal grading structure across geographically and technically diverse cohorts without dataset-specific recalibration, supporting robustness in real-world settings.

Importantly, our findings support an AI-assisted pathology paradigm rather than automation or replacement of expert judgment. MOSAIC does not seek to redefine grading criteria or supplant pathologist expertise, but instead provides standardized visual and quantitative evidence to guide attention, reduce search bias, and stabilize grading decisions. This framing is consistent with recent consensus in computational pathology that AI systems are most effective when deployed as decision-support tools integrated into routine diagnostic workflows rather than as autonomous classifiers.^1,10^

Several limitations should be acknowledged. Component-level pathologist scores were unavailable for all cohorts, restricting detailed concordance analyses outside TCGA-BRCA. Nuclear pleomorphism remains challenging for both human and AI-based assessment, highlighting the need for future refinement or alternative representations of nuclear atypia. Finally, while the Indian cohort demonstrated grading concordance, outcome data were not uniformly available across institutions.

### Clinical Implications

From a clinical perspective, MOSAIC addresses the inherent subjectivity of histologic grading, particularly at the low-intermediate boundaries that often complicate treatment decisions. By providing reproducible, explainable, and quantitative support within the existing Nottingham framework, MOSAIC standardizes reporting without requiring changes to established diagnostic criteria. This AI-assisted approach is especially valuable for resolving borderline cases, streamlining quality assurance, and ensuring consistency across institutions with varying levels of subspecialty expertise.

Beyond traditional grading, H&E-based analysis offers a potentially scalable alternative to gene-panel assays like Oncotype DX. While transcriptomic tests provide objective genomic risk assessment, they are frequently limited by high costs, long turnaround times, and the necessity of tissue destruction. By extracting high-dimensional features directly from standard slides, MOSAIC could maintain the objectivity of molecular testing while restoring critical spatial context. Although these H&E-based models require further prospective validation to match the clinical rigor of established assays, they represent a promising future direction for a more rapid, cost-effective, and non-destructive approach to precision oncology.

## Data Availability

All data produced in the present study are available upon reasonable request to the authors

## Supplementary Figures

**Supplementary Figure 1:**
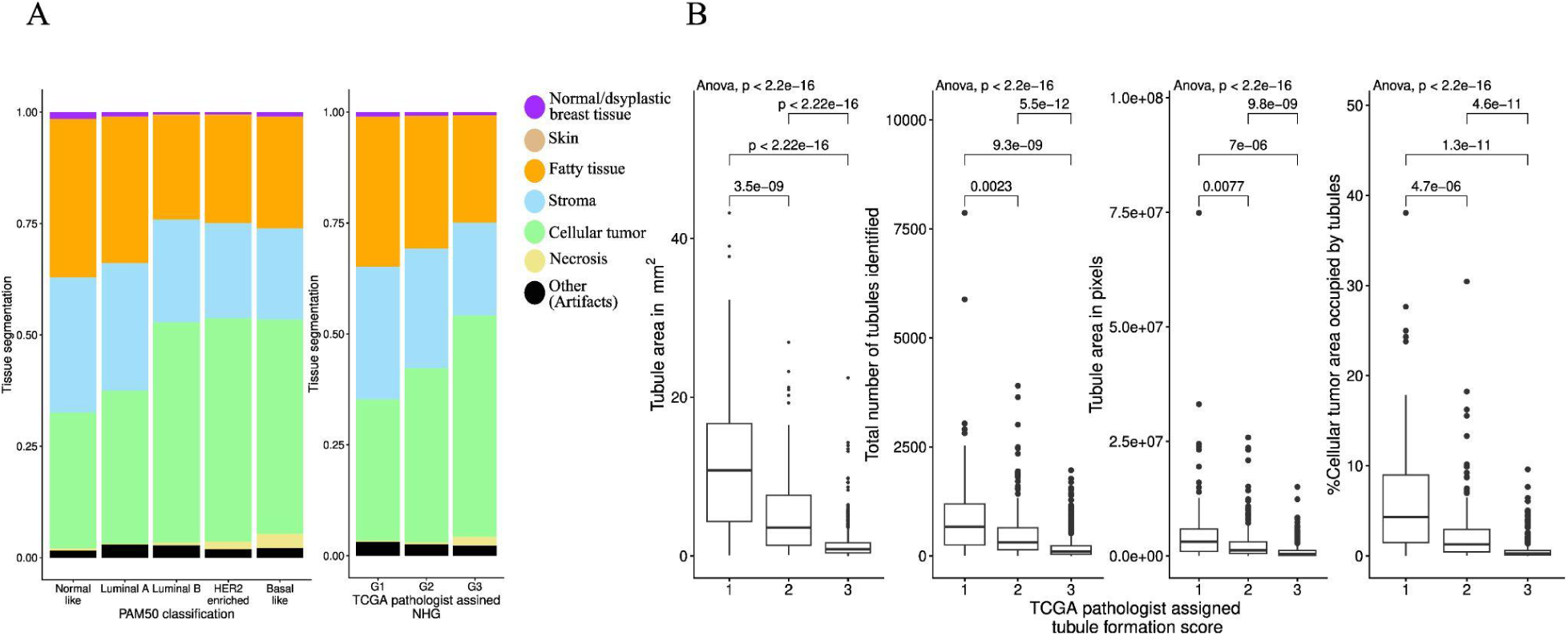
Tissue composition and quantitative tubule metrics across molecular subtype and pathologic scoring. (A) Left: Mean proportional area (%) of MOSAIC-derived tissue compartments stratified by PAM50 molecular subtype. Right: Mean proportional area (%) of tissue compartments stratified by TCGA-BRCA pathologist-assigned histologic grade. Values represent average WSI compartment percentages. (B) Distributions of quantitative tubule metrics stratified by TCGA pathologist-assigned tubule formation scores (1-3), including tubule density (tubules per mm²), raw tubule counts, total tubule area (pixels), and percentage of cellular tumor area occupied by tubules.

**Supplementary Figure 2:**
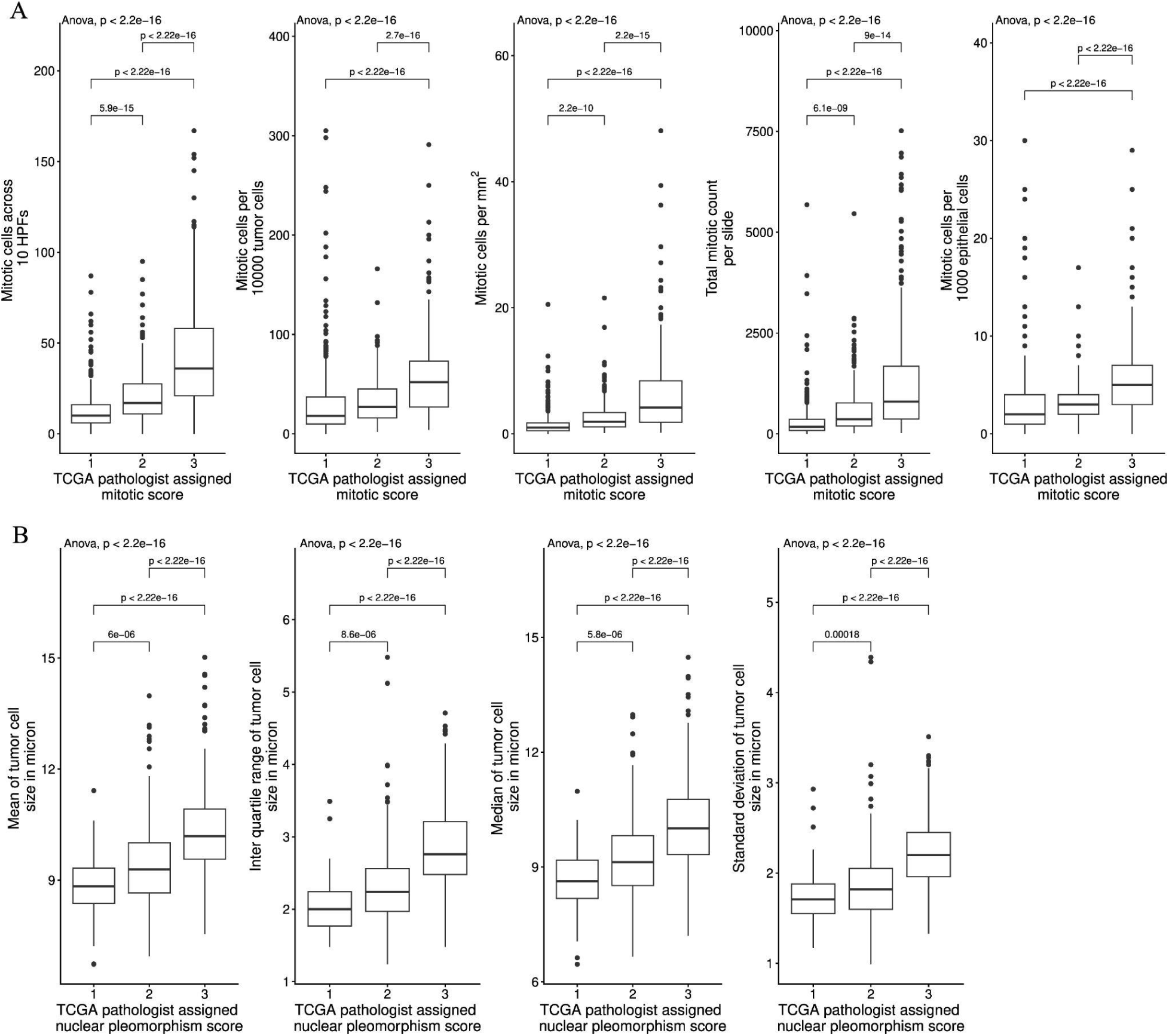
Association of automated mitotic and morphometric metrics with pathologist-assigned grading components. (A) Distribution of MOSAIC-derived mitotic metrics stratified by TCGA-BRCA pathologist-assigned mitotic activity score, including mitotic cells across 10 standardized HPFs (0.51 mm diameter each), mitotic cells per 10,000 tumor cells, mitotic cells per mm², total mitotic count per slide, and mitotic cells per 1,000 epithelial cells. (B) Distribution of tumor cell size metrics stratified by TCGA pathologist-assigned nuclear pleomorphism score, including mean, median, standard deviation, and interquartile range (IQR) of tumor cell size.

**Supplementary Figure 3:**
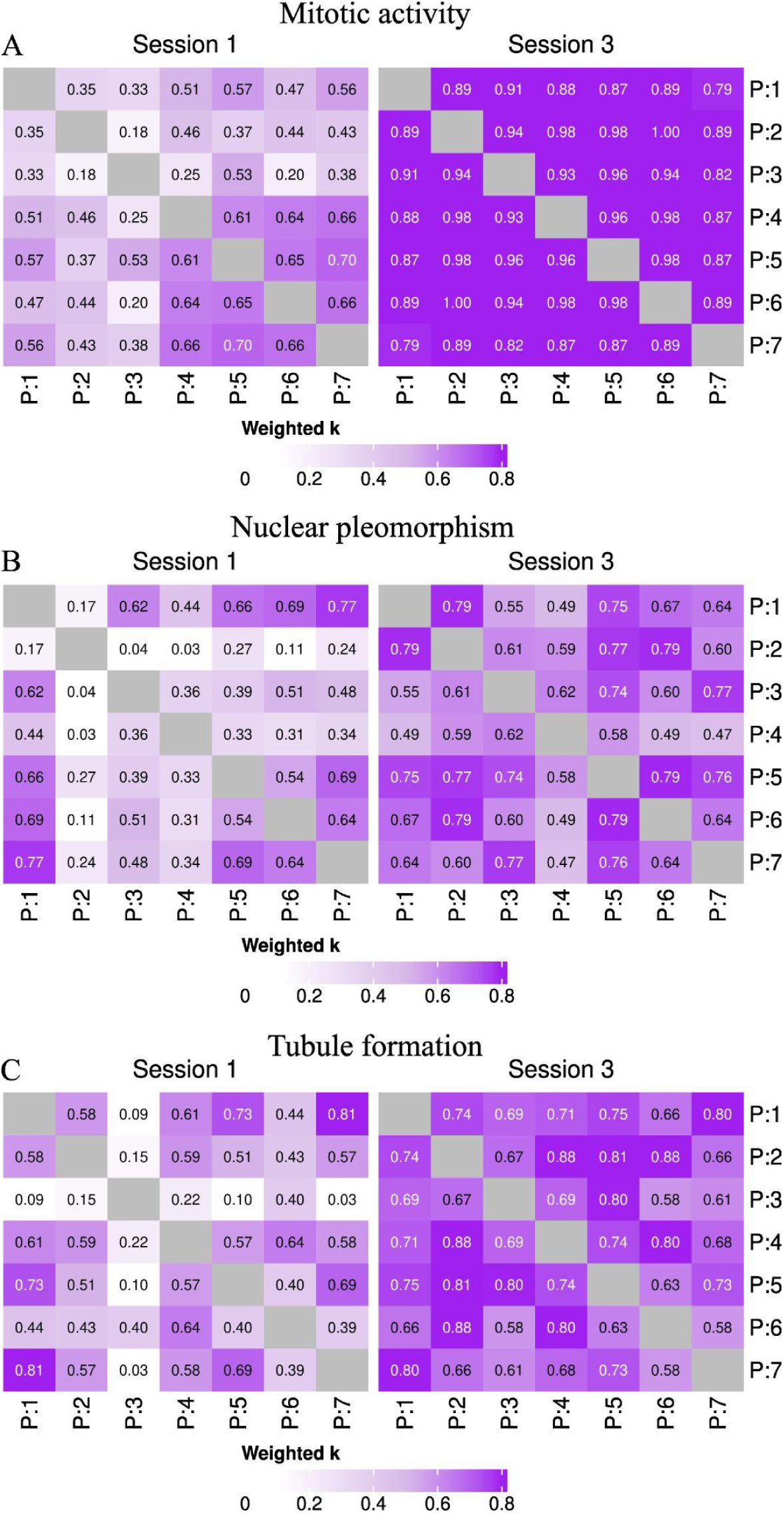
Inter-pathologist concordance in Phase 2 across grading components. Heatmaps display pairwise inter-pathologist agreement quantified using weighted Cohen’s kappa in Phase 2 Session 1 (n = 41 images) and Session 3 (n = 212 images). (A) Inter-pathologist concordance for mitotic activity. (B) Inter-pathologist concordance for nuclear pleomorphism. (C) Inter-pathologist concordance for tubule formation.

**Supplementary Figure 4:**
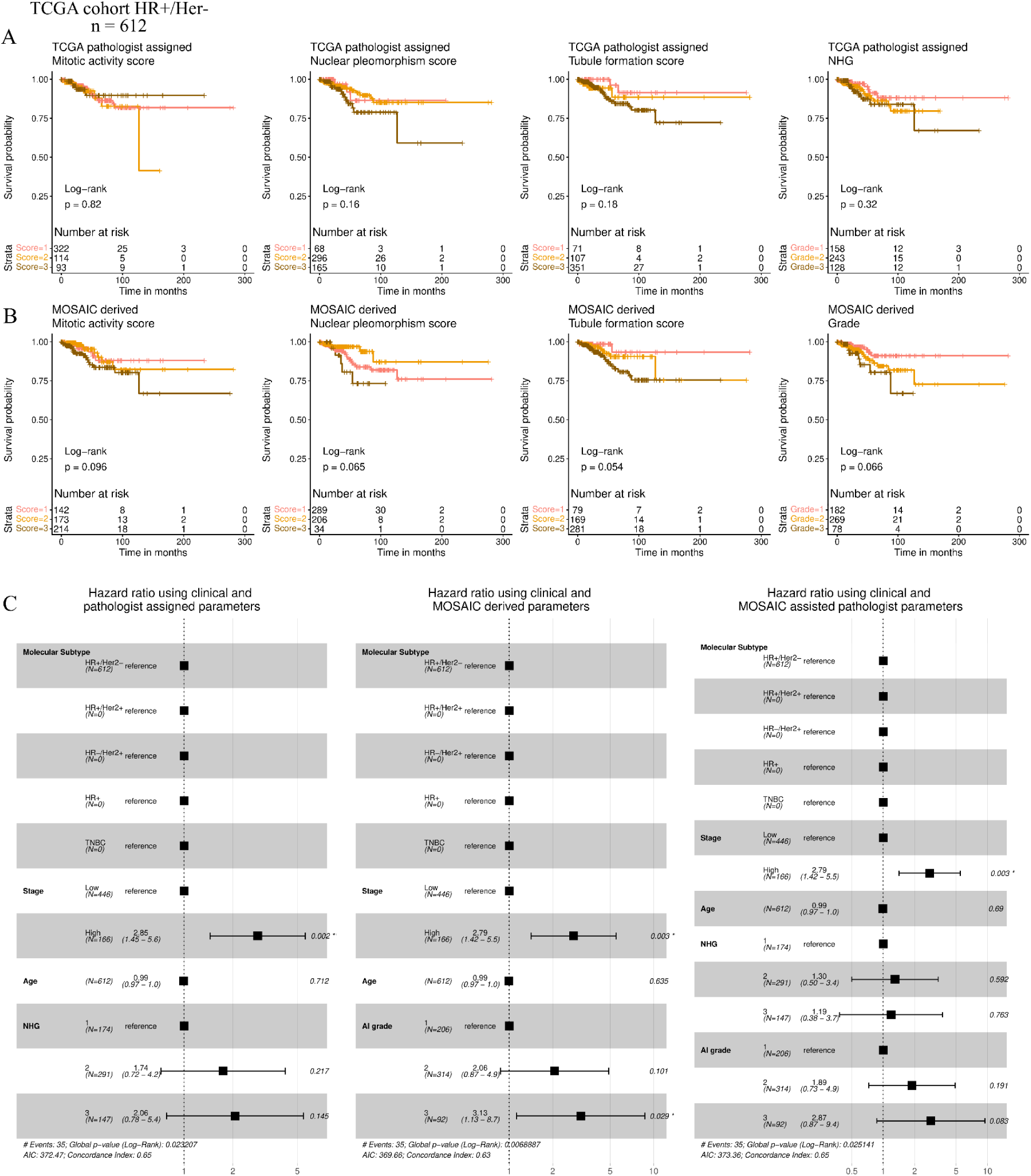
Prognostic stratification and multivariable survival analysis of MOSAIC-derived grading in the TCGA-BRCA HR+/Her2- cohort (n = 612). (A) Kaplan-Meier survival analysis stratified by pathologist-assigned mitotic activity, nuclear pleomorphism, tubule formation scores, and overall grade. (B) Kaplan-Meier survival analysis stratified by MOSAIC-derived mitotic activity, nuclear pleomorphism, tubule formation scores, and overall grade. (C) Multivariable Cox proportional hazards analysis incorporating clinical covariates, including age, stage, and molecular subtype. Forest plots show models using TCGA pathologist-assigned grade (left), MOSAIC-derived grade (middle), and a combined model including both grades (right).

**Supplementary Figure 5:**
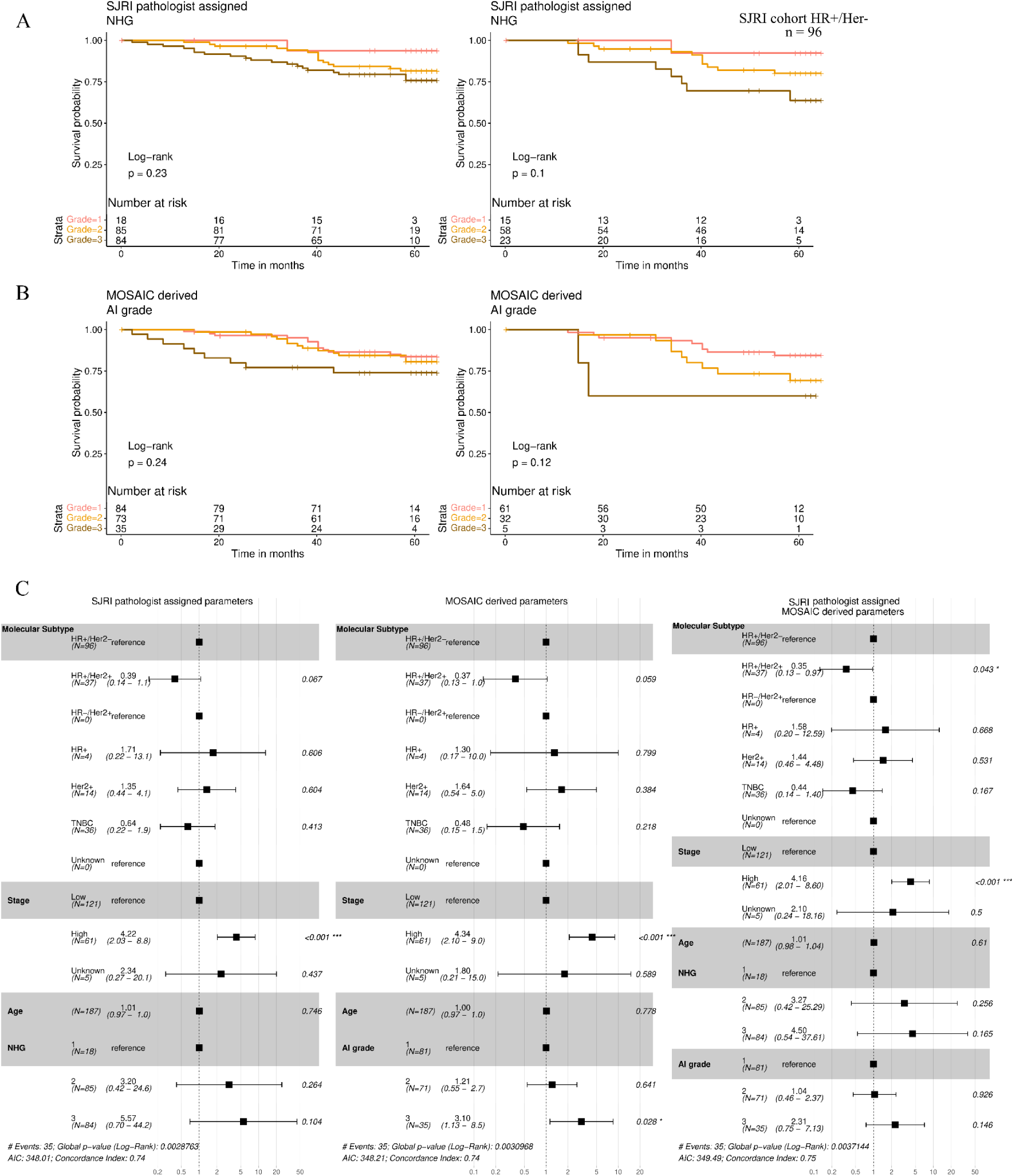
Prognostic stratification and multivariable survival analysis of MOSAIC-derived grading in the SJRI cohort (n = 192). (A) Kaplan-Meier survival analysis stratified by pathologist-assigned overall grade in the SJRI complete cohort (left) and the SJRI HR+/Her2- cohort (right). (B) Kaplan-Meier survival analysis stratified by MOSAIC-derived overall grade SJRI complete cohort (left) and SJRI HR+/Her2- cohort (right). (C) Multivariable Cox proportional hazards analysis incorporating clinical covariates, including age, stage, and molecular subtype. Forest plots show models using SJRI pathologist-assigned grade (left), MOSAIC-derived grade (middle), and a combined model including both grades (right).

